# Mapping the prevalence of molecular markers of *Plasmodium falciparum* artemisinin partial resistance in Africa: a spatial-temporal modelling study

**DOI:** 10.64898/2025.12.22.25342873

**Authors:** Neeva Wernsman Young, Cécile P. G. Meier-Scherling, Gina Cuomo-Dannenburg, George A. Tollefson, Sean V. Connelly, Jacob Marglous, Isabela Gerdes Gyuricza, Kelly Carey-Ewend, Ronald Kyong-Shin, Zachary R. Popkin-Hall, Ayalew Jejaw Zeleke, Deus S. Ishengoma, Abebe A. Fola, Alfred Simkin, Karamoko Niaré, Jonathan B. Parr, Melissa Conrad, Lucy C. Okell, Shazia Ruybal-Pesántez, Oliver J. Watson, Jonathan J. Juliano, Jeffrey A. Bailey, Robert Verity

**Affiliations:** Center for Computational Molecular Biology, Brown University, Providence, RI, USA; Department of Pathology and Laboratory Medicine, Brown University, Providence, RI, USA; MRC Centre for Global Infectious Disease Analysis, Imperial College, London, UK; Department of Microbiology and Immunology, Rega Institute, KU Leuven, Leuven, Belgium; MD-PhD Program, University of North Carolina, Chapel Hill, NC, USA; Institute for Global Health and Infectious Diseases, University of North Carolina, Chapel Hill, NC, USA; Curriculum in Genetics and Molecular Biology, University of North Carolina, Chapel Hill, NC, USA; National Institute of Biomedical Research, Kinshasa, Democratic Republic of the Congo; Department of Biology, Western Connecticut State University, Danbury, CT, USA; Department of Medical Parasitology, School of Biomedical and Laboratory Sciences, University of Gondar, Gondar, Ethiopia; Ifikara Health Institute, Ifikara, Tanzania; National Institute for Medical Research, Dar es Salaam, Tanzania; Division of Infectious Diseases, Department of Medicine, UNC School of Medicine, Chapel Hill, NC, USA; Department of Molecular Microbiology and Immunology, Johns Hopkins School of Public Health, Baltimore, MD, USA; Instituto de Microbiología, Universidad San Francisco de Quito, Quito, Ecuador; Department of Epidemiology, Gillings School of Global Public Health, UNC, Chapel Hill, NC, USA

**Keywords:** *Plasmodium falciparum*, malaria, antimalarial resistance, kelch, K13, MDR1, CRT, spatio-temporal modelling, Gaussian process, molecular epidemiology

## Abstract

**Background:** *Plasmodium falciparum kelch13* (*k13*) mutations in Africa signal emerging artemisinin partial resistance (ART-R), endangering malaria control by undermining artemisinin-based combination therapies (ACTs). Sparse surveillance obscures whether rising *k13* ART-R prevalence reflects local emergence or geographic expansion. We aimed to model and infer high-resolution spatial-temporal prevalence of *k13* ART-R and ACT partner-drug markers to inform public health policy.

**Methods:** We conducted a systematic literature review (PROSPERO-ID CRD42024593923) spanning the years 2014–2025, complementing existing data from WWARN, MalariaGEN Pf7, and the WHO Malaria Threats Map. This integrated dataset, comprising 3,806 distinct molecular epidemiology surveys and 182,071 genotyped samples, was harmonized using a standardized data schema. We applied a spatial-temporal Gaussian process model to estimate the continuous prevalence of WHO *k13* ART-R mutations, *mdr1* 86Y, and *crt* 76T.

**Findings:** ART-R increases were driven by distinct emergences of *k13* 561H in Rwanda, *k13* 675V in Uganda, and *k13* 622I in Ethiopia and Eritrea. From 2012 to 2024, predicted *k13* prevalence rose steadily in Northern Province, Uganda (1.81% per year) and Northern Province, Rwanda (3.49% per year), reaching 26.36% in Uganda and 39.44% in Rwanda by 2024. Modelling indicated a rapid transition from localized *k13* ART-R mutation emergence to entrenched regional hotspots centred on Uganda–Rwanda and the Ethiopia–Eritrea border. Partner drug amodiaquine marker *mdr1* 86Y is fading, but *crt* 76T remains prevalent in the Horn of Africa.

**Interpretation:** The rapid and multicentric expansion of *k13* ART-R mutations in East Africa threatens ACT efficacy, especially where ART-R *k13* and partner drug markers co-occur, mirroring early patterns observed before ACT failure in Southeast Asia. This study provides an updated *k13* ART-R mutation database and high-resolution resistance maps with uncertainty quantification, demonstrating a crucial need to prioritize targeted molecular surveillance across greater East Africa to safeguard ACT efficacy.

**Funding:** US National Institutes of Health.

**Research in Context:** *Evidence before this study:* We searched OVID MEDLINE and PubMed databases: Pubmed was searched from database inception to July 9th 2025, using the following search terms: (((((((plasmodium falciparum) OR (falciparum))) AND (Africa)) AND (((resistance) OR (kelch13) OR (pfkelch13) OR (k13) OR (crt) OR (pfcrt) OR (mdr1) OR (pfmdr1)))) AND (((Sequencing) OR (markers) OR (genotyping))). This search identified 922 publications, of which 116 meet our inclusion criteria. We extracted the prevalence of all WHO-validated and candidate *k13* mutations (441L, 446I, 449A, 458Y, 469F, 469Y, 476I, 481V, 493H, 515K, 527H, 537I, 537D, 538V, 539T, 543T, 553L, 561H, 568G, 574L, 580Y, 622I, 675V), *mdr1* 86Y, and *crt* 76T for these studies. In Africa, ART-R resistance mutations have been increasingly detected beginning in 2013-2015, particularly in East Africa (Rwanda, Uganda) and the Horn of Africa (Ethiopia, Eritrea). Therapeutic efficacy studies (TES) generally show high ACT efficacy across the continent; nevertheless, the presence of *k13* resistance mutations is viewed as a high-priority public health threat. Prior efforts to map these molecular markers have been hampered by sparse and heterogeneous sampling, resulting in prevalence maps that rely on simple aggregation with wide, uninformative uncertainty intervals, limiting utility for national programs seeking to identify true increases in resistance. Furthermore, sub-continental estimates have often been static or limited in their ability to model the spatial and temporal dynamics of distinct mutation lineages.

*Added value of this study:* This study provides the largest and most current harmonized dataset of *k13* ART-R and partner drug mutations in Africa with 182,071 samples and 3,806 surveys, integrating systematic literature review findings with major public databases (WWARN, MalariaGEN Pf7, and the WHO Malaria Threats Map). We used a spatial-temporal Gaussian process model to overcome the limitations of sparse sampling. This modelling approach enables the generation of continuous prevalence maps across space and time, simultaneously predicting prevalence in unsampled areas while rigorously quantifying the statistical uncertainty of those predictions. We provide the first contemporary, high-resolution maps that illustrate the concurrent selection dynamics of key partner drug markers (*mdr1* 86Y and *crt* 76T) alongside the expanding focus of ART-R.

*Implications of all the available evidence:* The dramatic rise in *k13* ART-R mutations, coupled with the steady geographic expansion identified by our model, confirms that ART-R is now firmly established in East Africa. The colocalization of high prevalence *k13* ART-R mutations in Uganda and Rwanda with near-fixation of markers related to partner drug tolerance creates a molecular resistance profile highly reminiscent of the conditions preceding widespread treatment failure in Southeast Asia. This suggests the need to more broadly shift away from reliance on artemether-lumefantrine and necessitates greater molecular surveillance in sub-Saharan Africa, prioritizing the identified high-prevalence regions (Uganda, Rwanda, Ethiopia, Eritrea) and neighboring countries. Our integrated approach provides the foundational, evidence-based tools needed by national malaria control programs (NMCPs) and the WHO to refine drug policy decisions before ART-R develops into clinical treatment failure.

## 2 Introduction

Despite the availability of effective frontline antimalarials, particularly artemisinin-based combination therapies (ACTs), malaria accounted for an estimated 265 million cases and 579,000 deaths in Africa in 2024, a burden that has remained largely unchanged in recent years.[1] Given that Africa bears the highest burden of malaria, the emergence and spread of artemisinin partial resistance (ART-R) in *Plasmodium falciparum* on the continent has the potential to stymie malaria control efforts and substantially increase morbidity and mortality if partner drug resistance were also to take hold.[2, 3, 4, 5, 6, 7, 8]

ART-R is defined clinically by delayed parasite clearance following treatment with an ACT.[9] This phenotype has been linked to mutations in the *kelch13* (*k13*) gene through *in vivo, in vitro* and reverse genetic engineering studies.[10, 11, 12, 13] The World Health Organization (WHO) uses this evidence to classify observed *k13* mutations as candidate or validated resistance polymorphisms, and recommends their use for tracking ART-R.[14] To date, more than 260 non-synonymous *k13* mutations have been reported, of which 23 are considered candidate or validated ART-R markers.[14] Understanding the distribution and spread of these molecular markers is an essential component of efforts to combat the emerging threat of antimalarial resistance globally.[15]

ART-R was first identified identified in western Cambodia in 2008 [9, 16] and was soon found to have spread to other parts of Cambodia, Thailand, Vietnam, Myanmar and Laos;[17] resistance to the artemisinin partner drug mefloquine, which emerged on the Cambodia-Thailand border in the 1990s, was already widespread when ACTs were introduced in this region.[18] Similarly, piperaquine resistance, mediated by *plasmepsin* 2/3 copy number variation and polymorphisms in the *P. falciparum* chloroquine resistance transporter (*crt*) gene, rapidly emerged after the introduction of dihydroartemisinin-piperaquine in 2008.[19, 20, 21] Co-occurrence of ART-R and partner drug resistance in Southeast Asia led to high rates of clinical failure of ACTs, reaching 50% by 2016.[22, 23, 24, 25] ART-R is now considered an urgent global public health priority, and a key component of the WHO’s global technical strategy for malaria 2016-2030. [26, 27]

Increasing reports of validated *k13* ART-R mutations in Africa raise concerns that resistance could follow a similar trajectory on a second continent. Initial modelling suggests no difference in the strength of selection for *k13* ART-R mutations in Africa vs. Asia [28]. However, therapeutic efficacy studies (TES) generally continue to show high ACT efficacy across Africa despite the emergence of ART-R [29, 30], with a few exceptions in Uganda, Democratic Republic of the Congo (DRC), Angola, and Burkina Faso [31, 32, 33, 34]. This is likely due to continued efficacy of the two most commonly used partner drugs, lumefantrine and amodiaquine, which are components of the widely-used ACTs artemether-lumefantrine (AL) and artesunate-amodiaquine (AS-AQ). However, reductions in artemisinin efficacy risk promoting rapid selection of partner drug resistance.[35, 36]

Estimating the burden and spatial distribution of antimalarial resistance markers at a continental scale remains challenging. Despite the rapid scale-up in sequencing of African isolates in recent years, sampling remains sparse and highly uneven, with large geographic areas lacking any molecular data and others represented by only a handful of samples.[37, 38, 39] In such settings, apparent patterns often rely on extrapolation from neighboring regions, despite differences in epidemiology and drug pressure that may limit comparability. Even where validated *k13* ART-R mutations have been detected, small sample sizes generate wide uncertainty intervals, making it difficult to distinguish true prevalence changes from random sampling error. This uncertainty has important implications for risk assessment, as policy decisions are frequently based on applying fixed thresholds without accounting for the confidence in the underlying data.

Statistical uncertainties are often compounded by issues around data harmonisation. Unlike routine epidemiological surveillance, molecular data exhibit substantial variability in sampling strategies, sequencing approaches, and down-stream bioinformatic processing. Data are also distributed over a wide range of public and private sources, and are stored in a range of file formats with limited inter-operability [40]. Even large-scale mapping initiatives, such as the Worldwide Antimalarial Resistance Network (WWARN) Molecular Surveyor [41] and the WHO Malaria Threats Map (WHO MTM) [42], present data in heterogeneous formats with unclear overlap between datasets.

Here, we address these gaps through an integrated approach that combines data aggregation, harmonisation, and statistical modelling. We conduct a systematic literature review of *P. falciparum k13* molecular data spanning 2014-2025, following the Preferred Reporting Items for Systematic Reviews and Meta-Analysis (PRISMA) guidelines. These data are merged with deduplicated datasets from multiple public sources, including WWARN [41], MalariaGEN Pf7 (Pf7) [43], and WHO MTM [42], producing the largest and most current dataset of *k13* polymorphisms in Africa to date. We further develop a standardised data schema for storing aggregated molecular data, allowing harmonisation over data sources and reducing opportunities for human error. Finally, we apply a spatial-temporal statistical model to the combined dataset to infer prevalence of mutations in unsampled areas while quantifying uncertainties. Our results provide a continent-wide view of the dynamics of *k13* ART-R mutations in Africa, offering critical insights for targeting molecular surveillance and informing national antimalarial treatment policies.

## 3 Methods

### Systematic Review

We undertook a systematic review of the literature to identify recent *P. falciparum* genotyping data not currently captured in WWARN [41], Pf7 [43], or WHO MTM [42]. The systematic review was conducted according to the PRISMA guidelines (PRISMA checklists, Supplementary Methods 2–3).[44] The review protocol was registered under PROSPERO ID CRD42024593923 [45]. We searched PubMed and MEDLINE databases from September 25, 2014 until 09 July 2025 for published studies reporting malaria resistance in Africa mediated via mutations in the *P. falciparum* genes *k13, mdr1*, or *crt* (Supplementary Methods 2). Briefly, we identified any study conducted in Africa that reported pre-treatment genotyping results for any *k13* mutation and/or *mdr1* N86Y and/or *crt* K76T, that are not already captured in an existing database (Supplementary Methods 3). In Covidence, two independent reviewers (from SC, NWY, CM-S, JM, GC-D, MC, JJJ, IGG, RK-S, KC-E) screened each title and abstract, and then subsequently assessed full-text for their eligibility for data extraction, with disagreements resolved by consensus between reviewers.[46].

In addition to the database search, we identified 13 privately held datasets that met the same eligibility criteria but were unpublished at the time of review. These datasets, which have all since been released as peer-reviewed articles or preprints, were extracted using the same procedures as the main review and were included in modelling. Given this blended approach, we refer to the combined dataset as the *augmented systematic review*.

### Data Extraction

Thirteen reviewers (NWY, CPGM-S, SVC, JM, GT, JJJ, KC-E, RK-S, IGG, MC, KN, AAF, AS) extracted the data from the augmented systematic review, collecting study overview information (country, PMID, publication year, etc.), site-specific information (site name, latitude and longitude, study design, etc.), and mutation prevalence per site for any *k13, mdr1* N86Y, and/or *crt* K76T mutation (Supplementary Methods 3). Data entry was manually checked by two reviewers to ensure the initial extraction did not contain any errors, and then data were uploaded to an internal server. Each study underwent preliminary data quality checks to ensure consistency of formatting of both prevalence data and study information.

### Data Integration and Harmonisation

Data from WWARN, WHO MTM and Pf7 were cleaned and deduplicated before combining with the augmented systematic review data (Supplementary Methods 5). Deduplication was via PubMed ID with ranked prioritisation: (i) augmented systematic review, (ii) WWARN, (iii) WHO MTM, (iv) Pf7. Wild type counts were imputed based on the reported portion of the *k13* gene sequenced (Supplementary Methods 7). Data harmonisation was achieved through the development of the STAVE software package (https://github.com/mrc-ide/STAVE), which provides a consistent format for aggregated genetic counts and their associated metadata through a relational database structure (Supplementary Methods 7). STAVE enforces a series of rigorous formatting checks, while also simplifying prevalence calculation through dedicated functions.

### Spatial-temporal Modelling

The prevalence of each molecular marker was modelled as a latent, continuous process varying smoothly across both space and time. Let *p*(*s, t*) denote the true prevalence at spatial location *s* ∈ ℝ ^2^ and time *t* ∈ ℝ. Observed counts *y*_*i*_ of mutant alleles at locations *s*_*i*_ and times *t*_*i*_ were assumed to follow a binomial distribution,

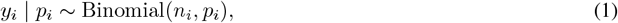

where *n*_*i*_ is the total number of successfully genotyped samples at this locus and *p*_*i*_ = *p*(*s*_*i*_, *t*_*i*_) is the latent prevalence at this location and time. To capture smooth variation, we defined a Gaussian process prior on the logit-transformed prevalence,

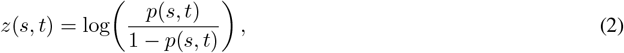

with separable spatial and temporal covariance structures. Spatial correlation was modelled using a squared exponential kernel parametrised by a length scale 𝓁. Temporal dependence was introduced through a Gaussian random-walk prior with variance parameter *τ* ^2^. Together, these assumptions define a spatial-temporal Gaussian process that allows gradual changes in prevalence while avoiding over-extrapolation when data are sparse. The spatial length scale 𝓁 was estimated by fitting a Gaussian model to the empirical spatial variogram, while the temporal variance parameter *τ* ^2^ was estimated by fitting a first-order random-walk model to the empirical temporal variogram (Supplementary Figures 5, 6).

Inference proceeds by estimating the latent surface *p*(*s, t*) from the binomial observations while accounting for uncertainty. For computational tractability with large datasets, we represented the Gaussian process using a reduced feature space, allowing approximate inference to obtain posterior median and uncertainty intervals for *p*(*s, t*). The fitted model yields continuous prevalence maps, enabling visualisation and quantification of resistance spread through space and time. Further information on the spatial-temporal model, the fitting approach, and the hyper-parameters used can be found in the Supplementary Methods 8-12.

Spatial–temporal modelling was restricted to *k13* ART-R mutations with sufficient data support, defined as detection in at least ten distinct prevalence surveys (441L, 449A, 469F, 469Y, 476I, 553L, 561H, 574L, 622I, and 675V), alongside *mdr1* 86Y and *crt* 76T. Posterior prevalence summaries are reported as the median of 1000 draws. We also calculated exceedance probabilities, which quantify the likelihood that prevalence exceeds a chosen threshold, in our case 5%. For each year, we identified pixels where the exceedance probability was greater than 80%, indicating high confidence that resistance was established. Summing the area of these pixels yielded an annual estimate of the geographic extent where resistance was confidently above the 5% threshold. Time series plots were produced by aggregating pixels to admin-1 regions and summarising using the median and 95% credible intervals over posterior draws.

## 4 Results

The systematic review identified 922 articles, of which 663 underwent screening after de-duplication (Figure 1). Screening removed 460 studies that did not match inclusion criteria. The remaining 203 studies underwent full review, which excluded an additional 87 studies that did not meet the selection criteria, resulting in 116 studies included in the final analysis (Figure 1). These were augmented with 13 privately held datasets, all since published as peer-reviewed articles or preprints (Supplemental Figure 2, Supplemental Methods 3). In total, the augmented systematic review identified 129 unique studies.

**Figure 1:**
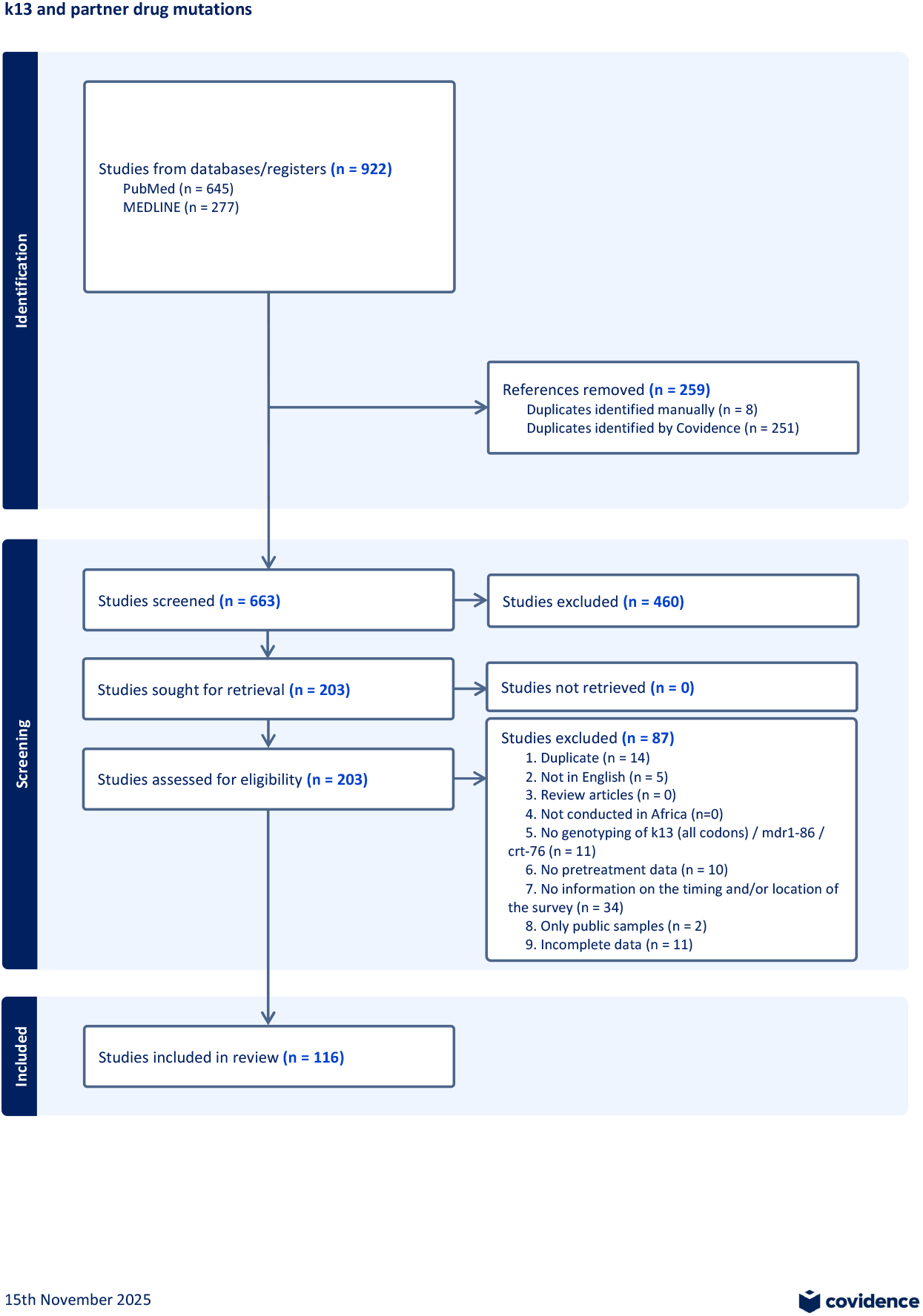
PRISMA Flowchart displaying the systematic literature review detailing our search from PubMed and Medline, the number of studies screened and the full texts retrieved.

Data from public repositories provided in an additional 450 studies after deduplication, comprising data from WWARN (363 studies), WHO MTM (34 studies) and MalariaGen Pf7 (53 studies). The final dataset included 93,553 samples sequenced at target *k13* positions and 182,071 samples sequenced for either *k13, crt* 76, or *mdr1* 86, drawn from 579 studies encompassing 3,806 distinct surveys across 47 African countries.

Eleven validated (446I, 469Y, 476I, 493H, 539T, 553L, 561H, 574L, 580Y, 622I, 675V) and seven candidate (441L, 449A, 469F, 515K, 527H, 538V, 568G) *k13* ART-R markers were observed over 17 African countries, identified across 72 studies spanning 1996-2024 (Figure 2; Supplementary Figures 3-4). Modelling indicates WHO-validated ART-R markers are currently mostly contained in East Africa, with three high prevalence regions associated with *k13* 561H spread in Rwanda, *k13* 675V and 469Y in Uganda and Rwanda, and *k13* 622I in Ethiopia and Eritrea. Initial emergence is spotty and restricted to small geographic regions, but overall *k13* ART-R mutations have expanded into regional hot-spots covering most of Uganda and Rwanda, and similarly at the border of Ethiopia, Eritrea and Sudan. This suggests that aggregated *k13* ART-R mutations are spreading outward from their initial foci (Figure 3A). Although current best estimates indicate a low prevalence of resistance mutations in surrounding countries (South Sudan, Kenya, Somalia), these are based on sparse and out-dated samples leading to high uncertainty. Hence, it is within the plausible posterior envelope that resistance has already spread over a much larger geographic range (Supplementary Figures 7-14).

**Figure 2:**
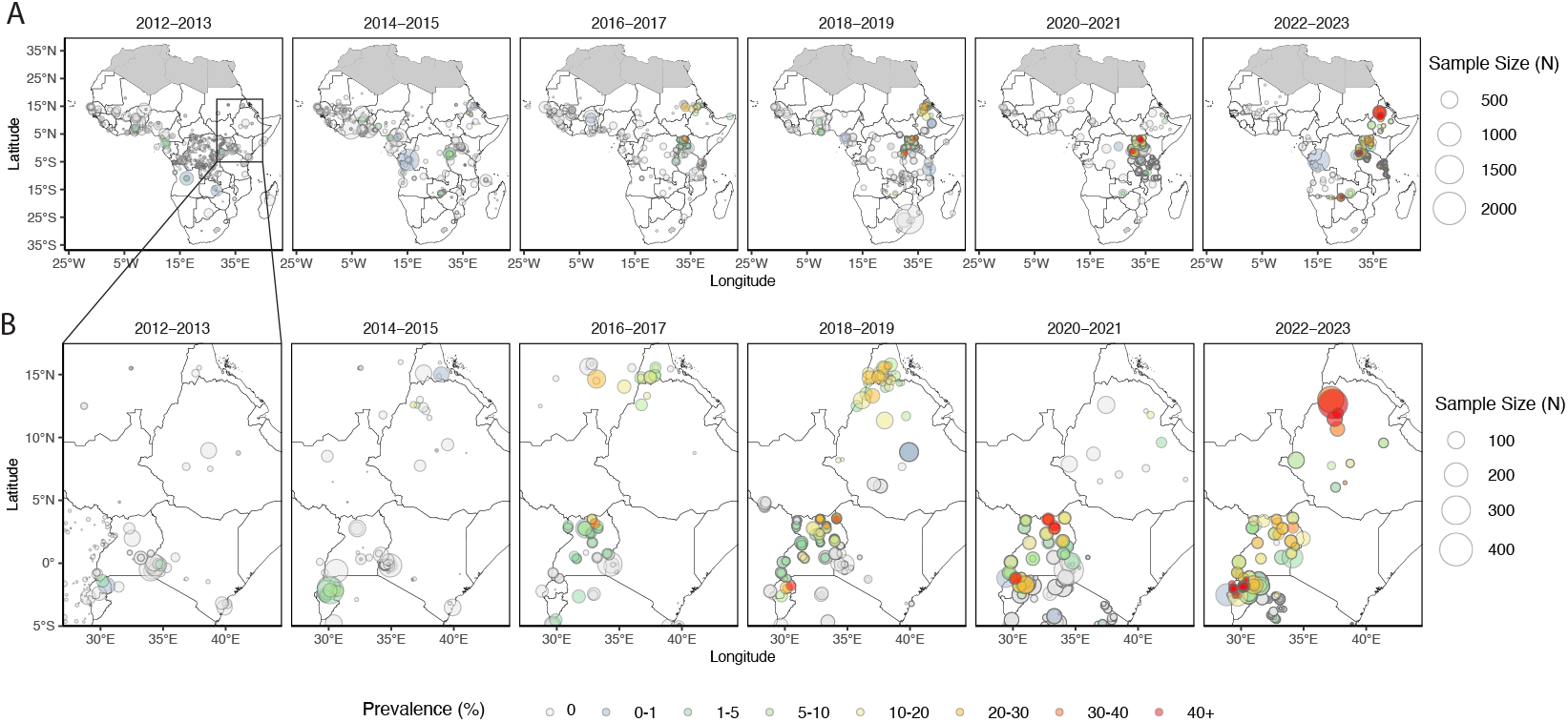
Observed prevalence of all WHO validated and candidate *k13* ART-R mutations combined across Africa, aggregated into two-year intervals. (A) Africa-wide maps showing prevalence by color and sample size by point size. (B) East Africa zoom of the same two-year–aggregated observations.

**Figure 3:**
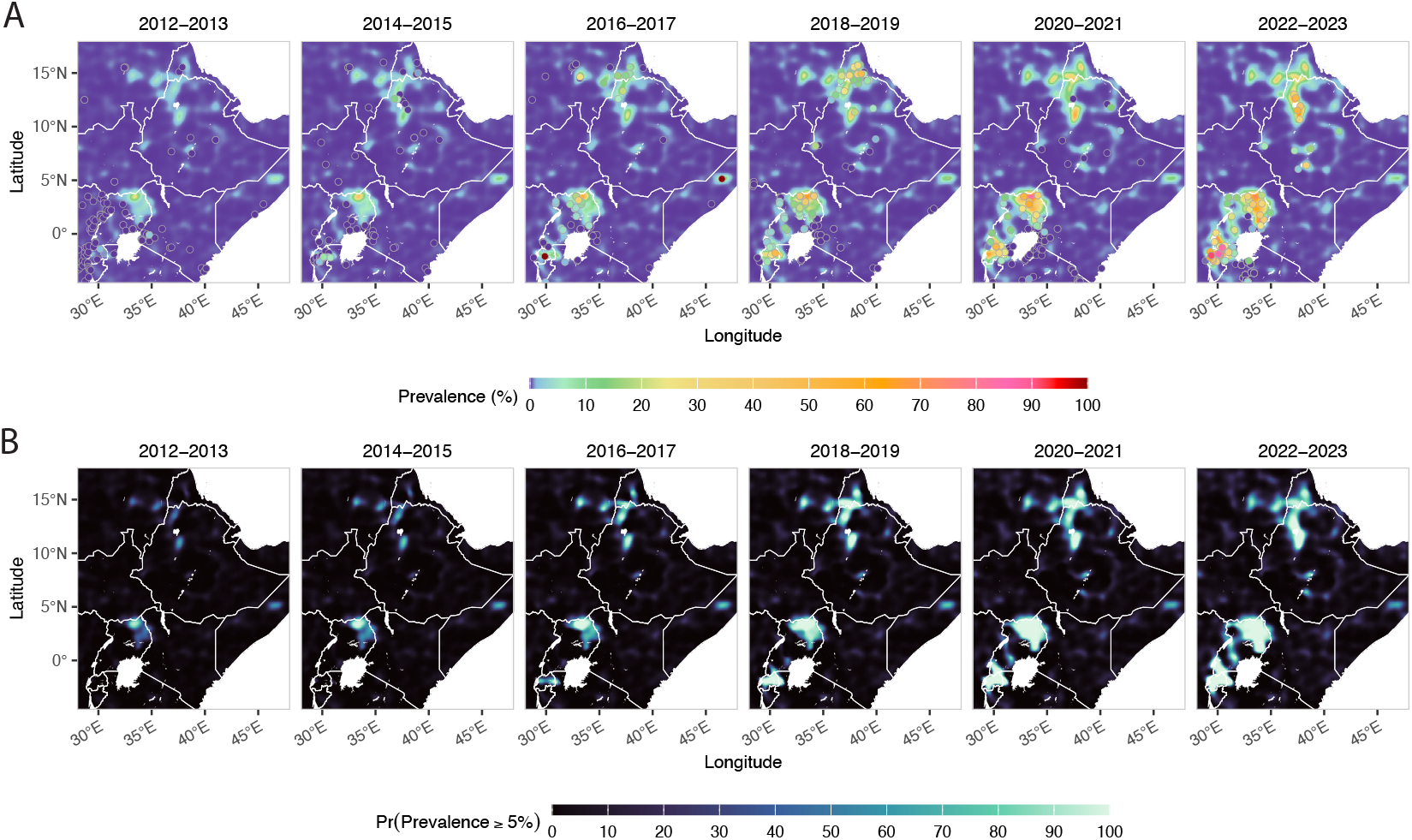
Posterior median *k13* mutations prevalence (A) and the posterior probability that true prevalence exceeds 5% (B), each averaged over two-year windows with observed data aggregated over the same periods, across East Africa from 2012–2023.

Exceedance probabilities highlight areas where the model is confident that the prevalence exceeds 5%. This is a composite measure that requires both high prevalence and high confidence (Figure 3B). Areas with high exceedance probability (>80%) in 2016 include Uganda and parts of Rwanda; these areas expand in subsequent years. The total area of high exceedance probability for all WHO-validated and candidate mutations increases from 5,879 km^2^ in 2013, roughly linearly to 188,622 km^2^ in 2024, with the largest area observed in 2022 at 208,954 km^2^. The overall pattern for all *k13* ART-R mutations combined is a steady increase despite staggered spread of individual mutations, hinting at a possible competitive interaction between strains (Supplementary Figure 15).

In 2012, the highest predicted prevalence of the 675V mutation was observed in Western Province, Rwanda (1.55%, 95% CI 0.34–7.27), increasing to 29.10% (95% CI 7.66–58.70) by 2024 in the same region. For the 561H mutation, the highest prevalence in 2012 occurred in Eastern Province, Rwanda (0.40%, 95% CI 0.15–1.14), rising to 36.00% (95% CI 7.00–80.40) by 2024, with the highest prevalence observed in Kigali City, Rwanda. 622I mutation predicted prevalence was highest in Aj Jazirah, Sudan in 2012 at 2.10% (95% CI 0.41–8.86), by 2024 the highest predicted prevalence came from Gash Barka, Ethiopia at 13.90% (95% CI 3.69–38.4) (Figure 4 and 5).

**Figure 4:**
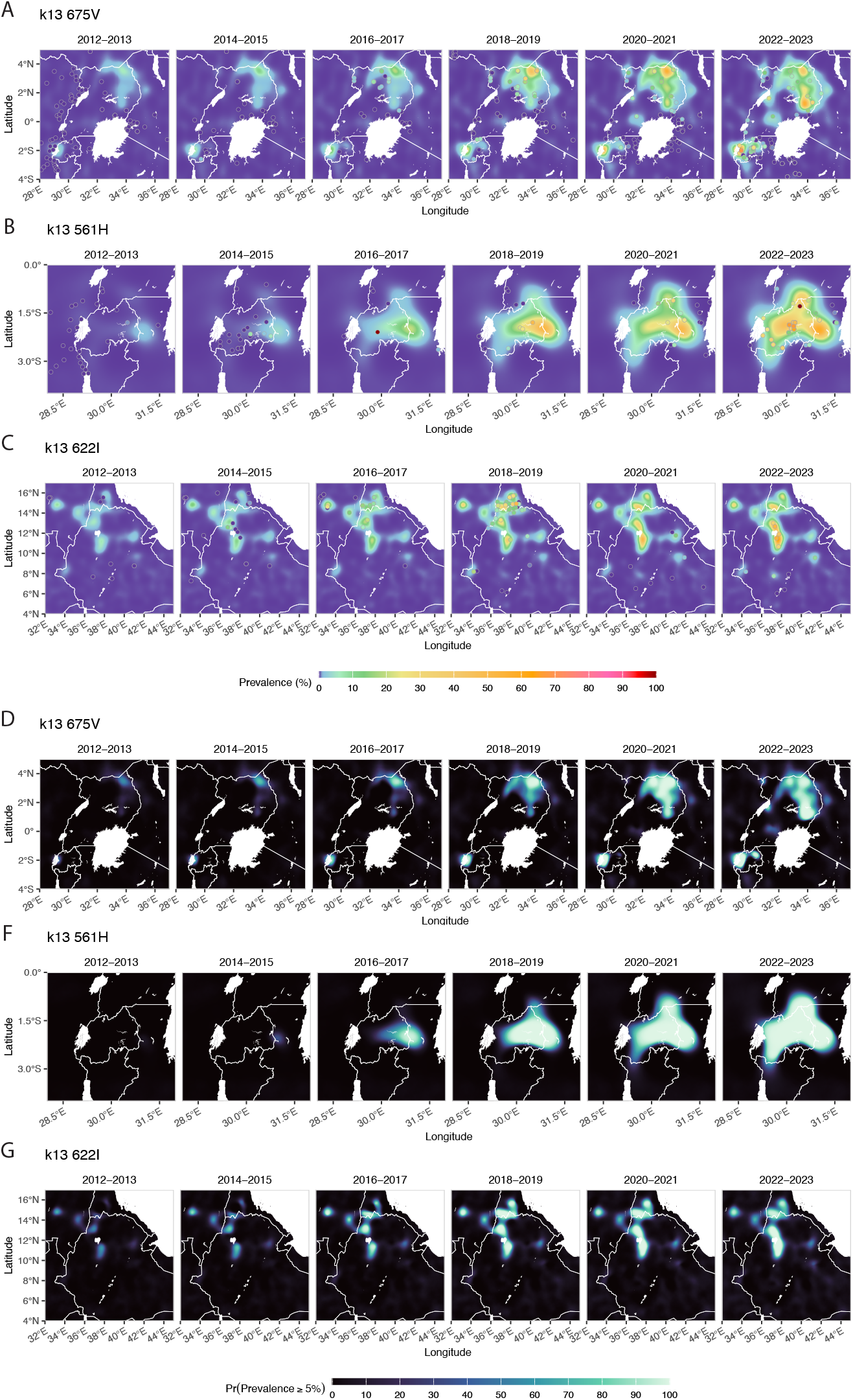
Posterior median prevalence for three *k13* mutations (675V, 561H, 622I) with observed data points (A–C) and the corresponding posterior probability that true prevalence exceeds 5% (D–F), each averaged over two-year windows with observed data aggregated over the same periods, across East Africa from 2013–2023.

**Figure 5:**
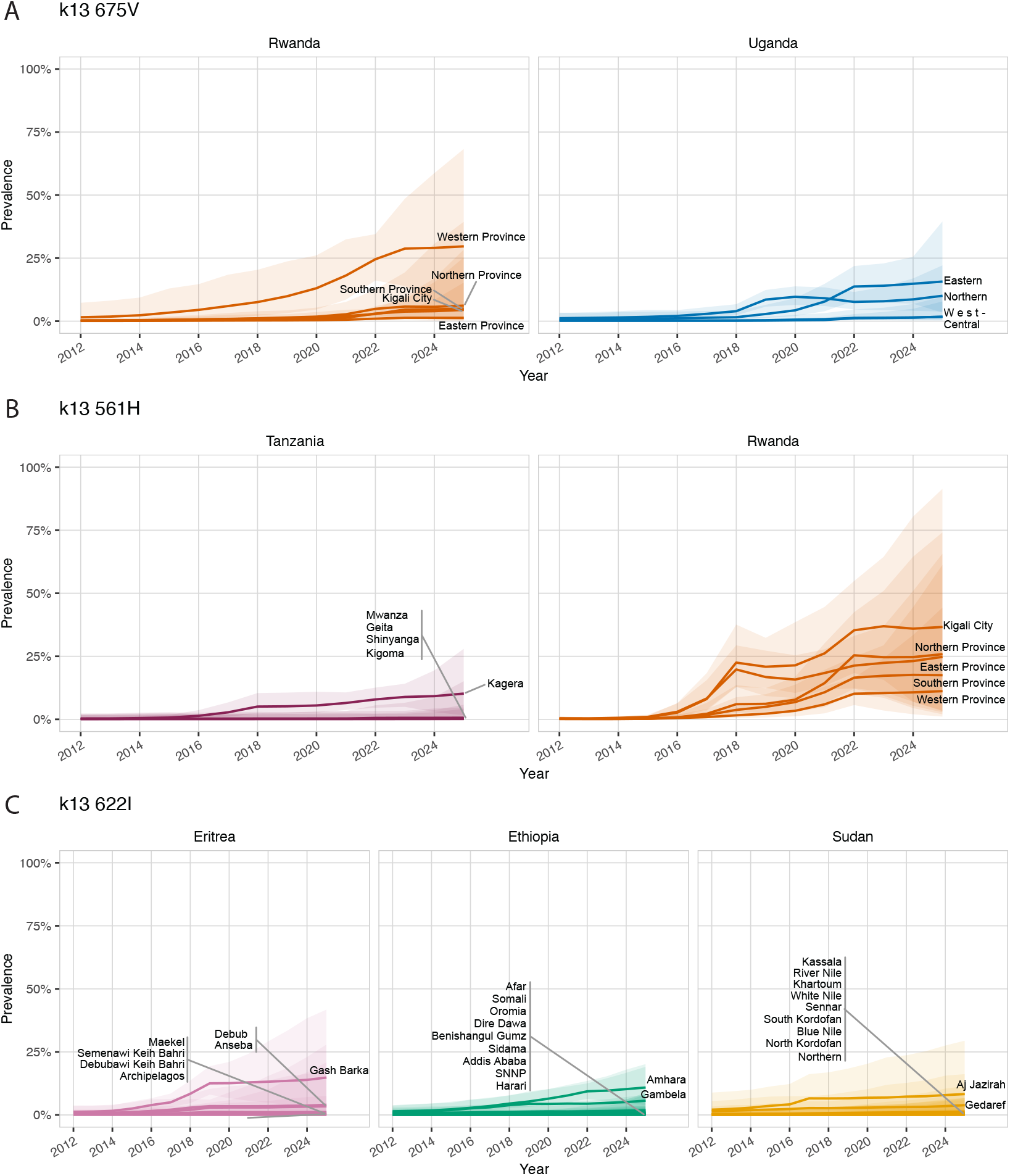
Time-series prevalence trends for *k13* 675V (A), *k13* 561H (B), and *k13* 622I (C) across Admin-1 regions in Rwanda, Uganda, Eritrea, Ethiopia, and Sudan where predicted prevalence exceeded 5%. Lines represent the posterior median predicted prevalence, and shaded ribbons indicate the 95% credible interval. These panels highlight when and where each mutation surpassed the 5% threshold and how trends evolved over time.

Other mutations show marked spatial and temporal heterogeneity. *k13* 469F first appeared at low prevalence in Rwanda in 2012 and later emerged at higher prevalence in southern Uganda in 2016, followed by wider regional spread in 2017. Variability in later years results in low exceedance probabilities and limited total area confidently above 5% prevalence. *k13* 469Y is mainly concentrated in northern Uganda and adjacent regions of South Sudan, Kenya, and the DRC. The area confidently above 5% prevalence rises steeply between 2019–2022 (from 8,084 to 43,726 km^2^), after which both prevalence and area estimates stabilise through 2022. Other candidate and validated mutations appear to have low overall prevalence with spotty and sporadic observations.

Figures 6 show the predicted prevalence of *crt* 76T (Panel A) and *mdr1* 86Y (Panel B), respectively. These mutations are associated with reduced susceptibility to amodiaquine ([47, 48, 49, 50]), the partner drug in AS-AQ, the second most common ACT in Africa. The prevalence of both mutations show significant spatial and temporal variation. High prevalence regions for *crt* 76T have historically been found throughout East, Central, and West Africa, but are now limited to the Horn of Africa (Figure 6; Supplemental Figures 16, 20) where they have remained highly prevalent due to ongoing chloroquine use to treat *Plasmodium vivax* malaria. The *mdr1* haplotype (N86, 184F, and D1246) has been linked to lumefanterine tolerance [51] which has followed the fading of 86Y and resurgence of N86 in countries with consistent AL usage (Supplemental Figures 17, 19), but does not alone make it predictive of partner drug failure.

**Figure 6:**
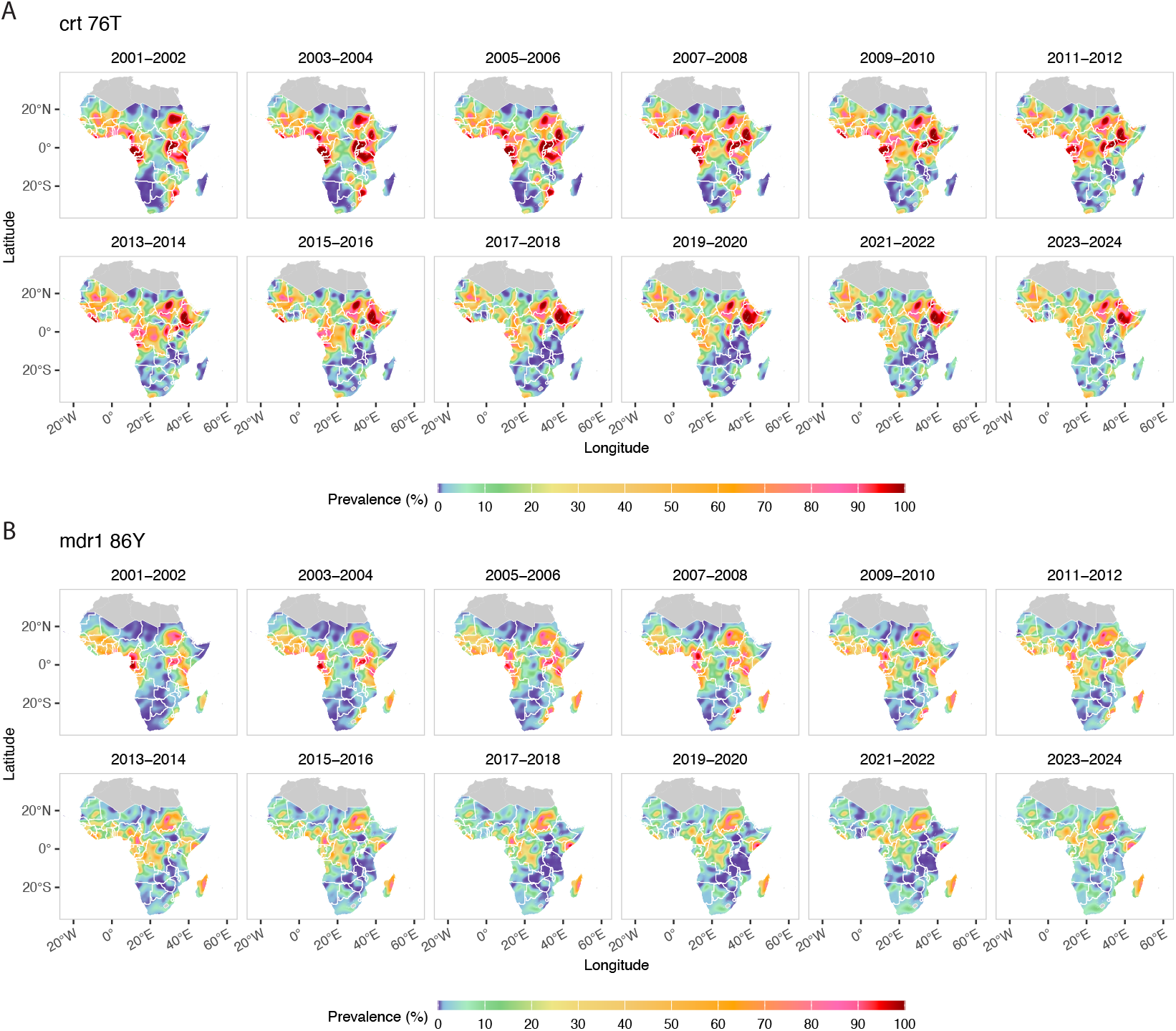
Posterior median *crt* 76T prevalence (A) and posterior median *mdr1* 86Y prevalence (B), each averaged over two-year windows with observed data aggregated over the same periods, across Africa from 2001–2024.

## 5 Discussion

Artemisinin partial resistance poses a serious threat to malaria control efforts in Africa. Molecular surveillance is central to tracking and responding to emerging resistance, as genetic data provide a scalable and timely indicator of resistance phenotypes that can be confirmed by therapeutic efficacy studies. However, past surveillance efforts have been predominately retrospective and hampered by challenges in data harmonisation. Unlike traditional epidemiological data, genetic data vary widely from targeted amplicon sequencing of a handful of loci to whole-genome sequencing. Data are distributed across multiple repositories with differing formats, objectives, and data standards [40]. This diversity is not a failure of the surveillance landscape but a reflection of the different purposes these resources were designed to serve, and as such, it is likely to persist. Effective continent-wide evaluation therefore requires integrative tools that can combine heterogeneous datasets rather than imposing uniformity upon them.

Raw molecular counts alone can give a misleading impression of resistance patterns. Apparent increases or absences often reflect variation in sampling and sequencing effort rather than true changes in prevalence. These issues are amplified by the uneven geographic distribution of sampling: large parts of Africa remain sparsely or entirely unsampled, and non-observance of detected mutations can reflect an absence of evidence rather than evidence of absence. Without careful interpretation, the pattern of sampling locations can be mistaken for biological signal, leading to over-interpretation of apparent hotspots or presumed safe zones. Statistical modelling is therefore essential for recovering underlying spatial–temporal trends and quantifying the uncertainty arising from data sparsity.

In this study, we assembled and harmonised the largest dataset of *k13, mdr1* N86Y, and *crt* K76T polymorphisms in Africa to date. A systematic review (PROSPERO ID CRD42024593923) was complemented by data extracted from three major repositories (WWARN, WHO MTM, and Pf7k) and further enriched with contributions from ongoing MMS efforts. Although this blended approach is not fully systematic, it reflects the practical and rapidly evolving nature of molecular surveillance in Africa, where new sequence data are produced at accelerating rates. Indeed, the fact that much of our most recent *k13* data was captured through the augmented systematic review rather than existing repositories reflects ongoing challenges in the timely deposition of sequence data during emerging crises (Supplemental Figure 2). By creating a unified format for aggregated genetic counts and metadata, our framework demonstrates a scalable approach for ongoing integration of molecular surveillance outputs by NMCPs, researchers, and global stakeholders. Realising the full value of such integrative frameworks will require data-sharing approaches that support timely cross-border inference while respecting national stewardship, attribution, and local ownership of molecular surveillance data.

Our analysis reveals a sustained and geographically expanding increase in *k13* ART-R mutations, which is no longer confined to isolated foci but represents an entrenched threat across East Africa. This is driven by multiple emergence events with distinct spatial and temporal trajectories. Despite substantial spatial overlap in their inferred ranges, these mutations are predominantly observed as distinct lineages, with co-occurrence within the same parasite populations remaining rare. This pattern suggests that resistance is spreading largely through the expansion of competing single-mutation bearing strains rather than through the accumulation of multiple *k13* variants, raising the possibility of fitness trade-offs that constrain multi-mutation genotypes. Understanding the complex relationship between genotype and phenotype will be crucial for interpreting the clinical significance and public health impact of molecular patterns.

Patterns of partner-drug tolerance further heighten concern. We observe substantial co-localisation of high *k13* ART-R mutation prevalence with markers of amodiaquine resistance (*mdr1* 86Y and *crt* 76T). *crt* 76T remains highly prevalent in East Africa and the Horn of Africa, while the *mdr1* N86, which is associated *in vitro* with AL usage [52], has risen to near fixation in countries relying on AL across much of sub-Saharan Africa. This molecular landscape is reminiscent of the conditions that preceded ACT failure in Southeast Asia, where simultaneous resistance to artemisinin and partner drugs led to widespread treatment failure. Although therapeutic efficacy studies in Africa still report high ACT efficacy, emerging evidence including increased *ex vivo* lumefantrine tolerance in Uganda [53], the identification of new haplotypes associated with both artemisinin resistance and lumefantrine tolerance [5], and reports of AL treatment failure in travellers all suggest that the buffer provided by partner drugs may be eroding [54, 31, 33, 32, 55, 56].

### Limitations

Despite the scale of our integrated dataset, substantial geographic gaps remain, and uncertainty intervals are wide in poorly-sampled regions such as South Sudan and Somalia. Small sample sizes limit precision, and heterogeneity in study design and genotyping methods may introduce bias despite harmonisation efforts. Our model presumes smooth spatial-temporal variation, which may not fully capture abrupt shifts due to local epidemiology, vector control, treatment policy, or population movement. Molecular markers, while powerful, do not directly measure clinical treatment failure, therefore continued therapeutic efficacy studies are essential. Finally, our analysis reflects data available up to July 2025; very recent or unpublished findings may not yet be captured.

Our spatial–temporal modelling framework provides actionable tools for NMCPs and global stakeholders to prioritise surveillance, anticipate risk, and refine drug policy before resistance leads to widespread treatment failure. Additionally, this framework is readily iterable, easily incorporating new data on novel and existing threats. Strengthening molecular surveillance, integrating genomic and therapeutic efficacy data, and evaluating alternative treatment strategies, including triple ACTs and new drug combinations, will be critical to mitigating the growing threat posed by the convergence of artemisinin partial resistance and partner-drug tolerance in Africa.

## Supporting information

Marker Prevalence Data

## Data Availability

All data and code for data cleaning, formatting and compilation (https://github.com/IDEELResearch/scrub), data harmonisation (https://github.com/mrc-ide/STAVE), raw data plotting (https://github.com/IDEELResearch/stitch), and modelling (https://github.com/mrc-ide/GRFFmap) are available on GitHub.

## 6 Acknowledgements

We thank UNC’s Winston House for hosting us during the Infectious Disease Epidemiology and Ecology Laboratory (IDEEL) 2025 London Hackathon, where this manuscript was conceived and work began on data abstraction and analysis. We thank an anonymous donor to the UNC Medical Foundation on behalf of Dr. Jonathan Juliano which funded the 2025 London IDEEL Hackathon.

## 7 Author Contributions

Conceptualisation (JBP, MC, OJW, JJJ, JAB, RV), Methodology (NWY, CPGMS, GCD, GAT, OJW, JAB, RV), Software (NWY, CPGMS, GCD, GAT, SVC, AS, SRP, OJW, JAB, RV), Validation (NWY, CPGMS, GCD, GAT, SVC, JM, RV), Formal Analysis (NWY, CPGMS, GCD, GAT, RV), Investigation (NWY, CPGMS, GCD, GAT, SVC, JM, IGG, KCE, RKS, ZRPH, AJZ, AAF, KN, JBP, MC, LCO, SRP, OJW, JJJ, JAB, RV), Resources (DSI, JJJ, JAB, RV), Data Curation (NWY, CPGMS, GCD, GAT, SVC, JM, LCO, OJW, JAB, RV), Writing – Original Draft (NWY, CPGMS, GCD, JBP, MC, JJJ, JAB, RV), Writing – Review and Editing (All), Visualisation (NWY, CPGMS, GCD, RV), Supervision (NWY, CPGMS, GCD, JJJ, JAB, RV), Project Administration (NWY, CPGMS, GCD, GAT, SVC, JJJ, JAB, RV), Funding Acquisition (JBP, MC, JJJ, JAB, RV).

## 11 Declaration of generative AI and AI-assisted technologies in the manuscript preparation process

During the preparation of this work the authors used ChatGPT 5.2 (v1.2025.343) in order to improve readability. After using this tool, the authors reviewed and edited the content as needed and take full responsibility for the content of the published article.

