## Supplemental text for "Mapping the prevalence of molecular markers of *Plasmodium falciparum* artemisinin partial resistance in Africa: a spatial-temporal modelling study"

December 22, 2025

### 1 Augmented Systematic Review

#### 1.1 Eligibility Criteria

We undertook a systematic review to identify new studies that satisfied the following eligibility criteria:

1. Full-text original research articles.
2. Written in English.
3. Published in peer-reviewed scientific journals.
4. Published between September 25, 2014, through July 9th 2025.
5. Samples were collected in Africa.
6. Genotyping results are reported for any *k13* mutation, *mdr1* N86Y polymorphism, and/or *crt* K76T polymorphism.
7. Information on the time of collection within a 1-year range (studies with aggregated data over multiple years that could not be separated were excluded)
8. Geographical location of participant sampling is provided to at least at Admin 1 level.
9. Samples collected came from citizens of the country, excluded travellers
10. Samples were reported with original sample size and specified how many samples were successfully genotyped.
11. Samples were collected prior to any treatment [ACT or Seasonal Malaria Chemoprevention (SMC)].
12. Samples were unique, originating from the group and were not taken from a public dataset.
13. Samples collected at day 0 of TES were sequenced agnostic to treatment outcomes.

##### 1.2 Study Selection

We conducted a systematic review with the following search terms in OVID MEDLINE and PubMed databases: Pubmed was searched from database inception to July 9th 2025 using the following search terms: (((((((plasmodium falciparum) OR (falciparum))) AND (Africa)) AND (((resistance) OR (kelch13) OR (pfkelch13) OR (k13) OR (crt) OR (pfcrt) OR (mdr1) OR (pfmdr1)))) AND (((Sequencing) OR (markers) OR (genotyping))).

All stages of the systematic review were conducted in the *Covidence* systematic review software [1]. Before studies were imported into Covidence, those that were already included in existing malaria molecular databases (WWARN [2], Pf7 [3], or WHO MTM [4]) were removed based on their PubMed IDs. This was in anticipation of data from existing molecular surveillance databases being combined with the systematic review data. Covidence software automatically identifies duplicate studies which are then excluded before title and abstract screening can commence.

From a team of 10 reviewers, two independent reviewers screened each title and abstract. Studies were kept at the title and abstract screening stage if they appeared to report pretreatment samples from Africa sequenced at *k13*, *mdr1* N86Y and/or *crt* K76T. If titles and abstracts were ambiguous, studies were retained for full text review. If the two reviewers disagreed about whether a study should be retained for full text review, a third reviewer would help them reach consensus. Covidence does not retain the reasons for exclusion at the title and abstract screening stage of the review, which is not required within the PRISMA guidelines.

For all studies retained after title and abstract screening, the full texts were obtained. From a team of 10 reviewers, two independent reviewers reviewed the full texts according to the following exclusion criteria. Studies were excluded under the first reason that they satisfy the exclusion criteria, with the criteria as follows:

1. Duplicate
2. Not in English
3. Review articles
4. Not conducted in Africa
5. No genotyping of *k13* (all codons) / *mdr1* 86 / *crt* 76
6. No pre-treatment data
7. No information on the timing and/or location of the survey
8. Secondary data
9. Incomplete data

###### 1.2.1 Structured Augmentation

In addition to the database search, we conducted a structured augmentation of the review to identify recent datasets that met the eligibility criteria but were privately held at the time of review. These datasets were extracted using the same procedure as the main review, which is detailed in the next section. All such private datasets have since been released as peer-reviewed articles or pre-prints.

In total, we extracted 129 studies: 116 from the systematic review and 13 from the structured augmentation.

##### 1.3 Data Extraction

Data extraction was preformed for the augmented systematic review within three separate tables: study-level, site-level and mutation-level.

- A study refers to an individual research effort that is linked to a publication reporting prevalence data. The study-level overview collects basic information about the study and the genotyping preformed.
- The site-level overview identifies the properties of the sampling location (e.g. country, lat/long) and the target population (e.g. age range, treatment status).
- The mutation-level overview records the prevalences of the genotyped markers for each site.

Example templates are given in Tables S1- S3.

Table S1: Study-level Overview Extraction Template, Formatting and Variable Names.

| **Data field** | **Expected data type** | **Variable name** | **Notes** |
|---|---|---|---|
| Countries in Study | Long Name | countries_covered | |
| Unique Study ID | Number_lowercasename_YYY | study_uid | Must match XLSX document name |
| Version Number | lowercase vXX | version | |
| Date Modified | YYYY-MM-DD | date_modified | |
| First Author Surname | Surname of first author, lowercase, no special characters | first_author_surname | |
| Publication Year | Year published YYYY, or "none" | publication_year | |
| PubMed ID | PMID only (not PMC), or blank | pmid | |
| Study URL | URL for publication or study site | study_url | |
| Publication Status | Peer-reviewed, preprint, unpublished, other | publication_status | |
| Genotype Method | Amplicon, WGS, MIP, Sanger, LDR-FMA, RFLP, etc. | genotype_method | |
| Genotype Panel | DR2, DR23KE, IBC, IBC2FULL, IBC2CORE, NA | genotype_panel | |
| *k13* minimum AA Position | Number between 1–726 or blank | *k13*_min | |
| *k13* maximum AA Position | Number between 1–726 or blank | *k13*_max | |
| *mdr1* N86Y Reported | yes or no | *mdr1*_N86Y | |
| *crt* K76T Reported | yes or no | *crt*_K76T | |
| Individual-level Data | tables, vcf, no, other | individual_level_available | |
| Haplotype Availability | *mdr1*, *crt*, DHPS, DHFR, Other | haplotypes_available | |
| Data Entry Author | Initials of data entry author | data_entry_author | |
| Data Processing Pipeline | Hand-entered, MIP, WGS, PfSMART | data_processing_pipeline | |
| Site-Level Overview Completion | yes or no | site_level_overview_complete | |
| Mutation Prevalence Completion | Wide (row-wise) or Long (column-wise) | mutation_prevalence_complete | |

Table S2: Site-level Overview Extraction Template, Formatting and Variable Names.

| Data field | Expected data type | Variable name | Notes |
|---|---|---|---|
| Site Unique ID | lowercase string | site_uid | |
| Site Name | Site Name from study; may match site uid | site_name | |
| Collection Location | Most specific location given (facility or town) | collection_location | |
| Country Name | Free text country Name | country | |
| ISO3C | three letter ISC03 code | iso3c | |
| Latitude (N) | signed decimal latitude | lat_n | |
| Longitude (E) | signed decimal longitude | lon_e | |
| Google Location | Map Search terms used for GPS lookup | google_lat_long_site | |
| Coordinate Source | origin of coordinates: manual, supplement, publication or author comments | | |
| Pretreatment Samples | Confirm pretreatment samples (yes or no) | pretreatment_samples | |
| Site Study Type | health_facility, community, TES, CPES, CSS, or Unknown | site_study_type | |
| Study Design Minimum age | if available, youngest age by number in years | study_design_age_min_years | |
| Study Design Max age | if available, oldest age by number in years | study_design_age_max_years | |
| Study Design Sex | if available, chose from both, male or female | study_design_sex | |

Table S3: Mutation-level Overview Extraction Template, Formatting and Variable Names.

| Data field | Expected data type | Variable name | Notes |
|---|---|---|---|
| Site Unique ID | lowercase string, must match one of the Site UIDs entered in the Site Level Overview | site_uid | |
| Substudy | Descriptor if data at site is broken down (day 0, day3, reinfection, recrudescence, untreated, etc) and (extracted or calculated to indicate if numbers were explicit or if Author has to calculate) | substudy | |
| Start Date | Most granular sample collection start date YYYY-MM-DD; can match end date if only one listed | date_start | |
| End Date | Most granular sample collection end data YYYY-MM-DD; can match start date if only one date | | |
| Genotyped Positions | Entered as Numerator / Denominator | Gene:Codon:Amino Acid (ex crt:76:T); haploytpes as Gene:Codon_Codon_Codon:AAA, Mixed Infections as Gene:Codon:A/A, Span of Wildtype as Gene:Codon-Codon:* | |

#### 2 Public Data Repositories: WWARN, WHO Malaria Threats Map, and MalariaGen Pf7

Data were extracted from the following public repositories:

- WWARN Artemisinin Molecular Surveyor and WWARN ACT Partner Drug Molecular Surveyor. Data downloaded 04 Dec 2023. Extracted *k13*, *crt* 76 and *mdr1* 86.
- WHO Malaria Threats Map. Data downloaded 02 Jan 2024. Extracted *k13* and *crt* 76 (*mdr1* data are present, but copy-number variation only so discarded).
- MalariaGen Pf7 drug resistance marker genotypes and associated study-level meta-data. Data downloaded 20 Sep 2024. Extracted *k13*, *crt* 76 and *mdr1* 86.

##### 2.1 WWARN Artemisinin Data Processing

WWARN data on *k13* ART-R mutations underwent the following processing steps. Numbers in brackets indicate the number of unique studies retained:

- Restrict to samples collected in Africa (107/211).
- Restrict to studies with valid PubMed IDs (103/107).
- PubMed IDs deduplicated against the augmented systematic review (94/103).
- Data cleaning, including fixing obvious data entry mistakes by reference to the source paper. In some cases the issues could not be resolved by reference to the source, in which case these studies were excluded (92/94).
- WHO candidate or validated loci must be covered by sequencing range (91/92).

##### 2.2 WWARN Partner Drug Data Processing

WWARN data on *crt* 76 and *mdr1* 86 mutations underwent the following processing steps:

- Restrict to samples collected in Africa (315/539).
- Restrict to studies with valid PubMed IDs (311/315).
- PubMed IDs deduplicated against the augmented systematic review (310/311).
- Data cleaning, including fixing obvious data entry mistakes by reference to the source paper. In some cases the issues could not be resolved by reference to the source, in which case these studies were excluded (290/310).
- During cleaning of WWARN *k13* data, we identified 5 studies where partner drug data are also available but were missing from WWARN Partner Drug dataset. We added these studies in, hence total studies increased by 5 (295).

The combination of WWARN artemisinin and partner drug data represents 363 distinct studies (91 artemisinin + 295 partner drug - 23 overlap).

##### 2.3 WHO Malaria Threats Map Data Processing

WHO Malaria Threats Map data on *k13* and *crt* 76 underwent the following processing steps:

- Restrict to samples collected in Africa (155/290).
- Restrict to studies with valid PubMed IDs (112/155).
- PubMed IDs deduplicated against the augmented systematic review (94/112).
- PubMed IDs deduplicated against WWARN (24/94).
- Data cleaning, including fixing obvious data entry mistakes by reference to the source paper (all studies retained).
- WHO candidate or validated loci must be covered by sequencing range (23/24).

##### 2.4 MalariaGen Pf7 Data Processing

MalariaGen Pf7 data on *k13* and *crt* 76 and *mdr1* underwent the following processing steps:

- Merge study meta-data with drug resistance data.
- Restrict to samples collected in Africa (53/80).

No further filtering of Pf7 was required. These studies do not have associated PubMed IDs, and therefore were not deduplicated against other databases.

##### 2.5 Combined Data Metrics

The combined dataset comprises the following total number of samples sequenced at each position:

- *k13* 561: 87,400 samples.
- *k13* 622: 83,069 samples.
- *k13* 675: 82,288 samples.
- *k13* any WHO candidate or validated position: 93,553 samples.
- *crt* 76: 114,555 samples.
- *mdr1* 86: 103,792 samples.
- *k13* any WHO candidate or validated position, or *crt* 76 or *mdr1* 86: 182,071 samples.

Note, these numbers are after imputation of wild type (see next section) and includes surveys that found only wild type.

#### 3 Imputation of Wild Type Counts

Across both the augmented systematic review and public data repositories, *k13* mutations were typically reported as individual deviations from the wild type. For example, a study might report that 2 out of 50 samples carried the A675V mutation. However, sequencing often covers a broader set of codons — frequently the entire *k13* propeller domain — meaning these data imply that wild type was observed at all other sequenced positions. It is important that we capture this wild type information to avoid ascertainment bias, as without it we would only have prevalence data when a mutant was observed, and would be missing a large proportion of zero prevalence points.

It is rarely possible to unambiguously determine the correct denominator at each wild type position, as sequencing coverage tends to vary across the gene and is rarely reported for invariant sites. We chose to impute wild type denominators using a conservative strategy. For each study, across both the augmented systematic review and public repositories, we first extracted the range of *k13* positions that were reported as covered by sequencing. If any *k13* mutation was observed within a given survey (defined by site and collection period), the wild type denominator was set to the *smallest* reported denominator across all *k13* positions in that survey. If no *k13* mutations were detected, but counts were reported for *crt* 76 or *mdr1* 86, the wild type denominator was set to the smallest denominator observed at those loci. This approach avoids overstating the strength of negative evidence while allowing zero counts to be explicitly captured.

#### 4 Data Harmonization in STAVE

##### 4.1 Objectives

Once data were extracted and cleaned, they were harmonized through a newly developed software package called STAVE, which stands for Spatial-Temporal Aggregated Variant Encoding.

STAVE has three focused capabilities:

1. **Compact storage of aggregated variants**. Encodes numerator/denominator amino-acid variation (single codons or multi-locus haplotypes) into a concise string format tailored for drug-resistance markers.
2. **Space–time linkage**. Associates each genetic observation with a precise geographic coordinate (latitude and longitude) and collection time (day), reducing ambiguity and potential for human error.
3. **Prevalence calculation**. Computes variant prevalence from encoded strings, correctly handling partial haplotypes, mixed calls, and other common aggregation complexities.

STAVE requires input data in a strict format, and includes a series of basic checks that will reject data if not conforming to this format. All four data streams described above were converted to STAVE format and imported into a single data object. Once in this format they could be easily manipulated and processed.

##### 4.2 Relational Structure

STAVE organizes data into three linked tables:

1. **Studies:** records the data source (e.g., publication) and provides the top-level relational identifier.
2. **Surveys:** represents each discrete sampling event, storing the associated space–time metadata (latitude, longitude, collection date). A single study may contain multiple surveys.
3. **Counts:** stores the aggregated genetic observations, including the variant identifier (gene, locus, mutation) and the corresponding numerator and denominator.

These tables are connected by two sets of relational keys: Studies → Surveys, and Surveys → Counts, ensuring that every genetic observation is linked to its sampling event and original source without redundancy.

Further details on the format of these tables, and how this facilitates prevalence calculation, can be found in the online documentation (https://mrc-ide.github.io/STAVE/).

##### 4.3 Versioning and Availability

STAVE is an actively maintained and developed piece of software. The version used in this project is v1.1.0, released 17 April 2025. STAVE is open-source and released under the MIT license.

#### 5 Spatial-temporal model

##### 5.1 Model Overview

We model the spatial-temporal prevalence $p(s,t) \in [0,1]$ of a chosen resistance marker on a two-dimensional spatial domain $s \in \mathcal{S} \subset \mathbb{R}^2$ and a set of discrete time points $t = 1, \ldots, T$.

The data are observed at a subset of locations $s_i$ and times $t_i$ for $i = 1 : N$. The prevalence at these locations and times can be written as $p_i = p(s_i, t_i)$. Data are assumed to be binomial counts around this latent prevalence:

$$y_i \mid p_i \sim \text{Binomial}(n_i,\, p_i) \tag{1}$$

where $y_i$ is the number of samples found to carry the mutation, and $n_i$ is the total number of samples successfully sequenced at this locus.

We assume a latent Gaussian spatial-temporal process for the logit-transformed prevalence:

$$z(s,t) = \text{logit}\, p(s,t) = \log\left(\frac{p(s,t)}{1-p(s,t)}\right). \tag{2}$$

Our goal is to infer the latent spatial-temporal surface $p(s,t)$ given the observed data $\mathbf{y} = (y_1, \ldots, y_N)$. To make inference computationally tractable, we employ a series of approximations and algorithmic techniques that exploit the structure of the model.

##### 5.2 Latent process model (state-space formulation)

###### 5.2.1 Random Fourier Feature (RFF) spatial representation

We assume independent covariance structures in space and time. For the spatial kernel, we assume a squared exponential (SE) form with length-scale $\ell$:

$$k_{\text{space}}(s,s') = \exp\left(-\frac{|s-s'|^2}{2\ell^2}\right). \tag{3}$$

Note that the overall variance term commonly seen in the SE kernel is omitted as it is absorbed in the temporal covariance term. Direct inference with this kernel has computational cost that scales with $\mathcal{O}(N^3)$ due to the matrix inversion of the $N \times N$ kernel matrix, making it infeasible for large datasets. To enable scalable computation, we approximate the Gaussian process using the *Random Fourier Feature* (RFF) method [5], which provides an explicit, low-dimensional feature representation of the kernel.

Specifically, we approximate the SE kernel using its spectral representation, as given by Bochner's theorem [6, 7], which states that any stationary kernel can be expressed as the Fourier transform of a positive spectral density function. For the SE kernel, this spectral density is Gaussian with variance $\ell^{-2}$. Accordingly, we draw $D$ random frequencies:

$$\omega_j \sim \mathcal{N}(0, \ell^{-2} I_2), \quad j = 1, \ldots, D, \tag{4}$$

and we use these random frequencies to define the feature map:

$$\phi(s) = \frac{1}{\sqrt{D}} \left[\cos(\omega_1^\top s), \ldots, \cos(\omega_D^\top s)\ ,\ \sin(\omega_1^\top s), \ldots, \sin(\omega_D^\top s)\right]^\top \in \mathbb{R}^{2D}. \tag{5}$$

The logit-prevalence field is represented as a linear combination of features:

$$z(s,t) = \mu + \phi(s)^\top \beta_t, \tag{6}$$

where $\beta_t \in \mathbb{R}^{2D}$ is the vector of feature coefficients at time $t$. $\mu$ is an intercept that we fix to logit($p_{\text{init}}$), the assumed initial prevalence on the logit scale.

###### 5.2.2 Temporal evolution (state dynamics)

In contrast to the spatial covariance, which admits a stationary kernel and thus a spectral (RFF) representation, we model temporal dependence directly in the latent feature coefficients $\beta_t$. We treat time as discrete and represent temporal evolution through a simple state-space (Markov) model. This is computationally efficient because the Markov property means that computation scales linearly with the number of time points ($O(T)$), avoiding the cubic scaling ($O(T^3)$) associated with full temporal GPs.

Specifically, we assume that the coefficients $\beta_t \in \mathbb{R}^{2D}$ evolve smoothly over time according to a Gaussian random walk — that is, a first-order autoregressive process with unit persistence:

$$\beta_t = \beta_{t-1} + \xi_t, \qquad \xi_t \sim \mathcal{N}(0, \tau^2 I_{2D}). \tag{7}$$

Here, $\tau^2$ controls the degree of temporal smoothness: smaller values enforce stronger persistence over time (i.e., slower temporal variation), while larger values allow more rapid temporal change.

Unlike the spatial component, there is no stationary temporal kernel $k_{\text{time}}(t, t')$ with a known spectral density from which RFFs could be drawn. Instead, this Markovian random-walk formulation plays an analogous role by inducing temporal covariance:

$$\text{Cov}(\beta_t, \beta_{t'}) = \tau^2 \min(t, t') I_{2D}, \tag{8}$$

which corresponds to a Brownian motion prior on the latent coefficients and hence a temporally smooth but nonstationary evolution of the spatial field.

The initial state is taken to be deterministic ($\beta_1 = 0$) reflecting the assumption of zero uncertainty in the initial prevalence $p_{\text{init}}$. This allows us to "pin" the starting prevalence to an appropriate small value that captures the situation before the widespread introduction of resistance mutations.

This approach defines a linear Gaussian state-space model on the coefficients $\beta_t$, which translates into a spatial-temporal GP prior on $z(s,t)$.

##### 5.3 Observation model (Binomial likelihood)

As mentioned above, let $y_i$ denote the number of observed mutant alleles at spatial location $s_i \in \mathbb{R}^2$ and sampling time $t_i \in \{1, \ldots, T\}$, taken from $n_i$ total samples. Each observation is modeled as a binomial draw:

$$y_i \mid p_i \sim \text{Binomial}(n_i, p_i), \tag{9}$$

where $p_i = p(s_i, t_i)$ is the true prevalence at the sampling location and time. We link the prevalence to the latent logit-scale field through the RFF representation:

$$\text{logit}(p_i) = z(s_i, t_i) = \mu + \phi(s_i)^\top \beta_{t_i}, \tag{10}$$

where $\phi(s_i) \in \mathbb{R}^{2D}$ is the feature vector at location $s_i$, and $\beta_{t_i}$ are the feature coefficients corresponding to the sampling time $t_i$.

Assuming conditional independence of observations given the latent process $\{\beta_t\}_{t=1}^T$, the joint probability of all observed data is:

$$p(\mathbf{y} \mid \{\beta_t\}_{t=1}^T) = \prod_{i=1}^{N} \binom{n_i}{y_i} p_i^{y_i} (1 - p_i)^{n_i - y_i}, \qquad p_i = \text{logit}^{-1}\big(\mu + \phi(s_i)^\top \beta_{t_i}\big). \tag{11}$$

This likelihood is non-Gaussian, preventing closed-form Kalman updates – a set of recursive equations used to optimally estimate the unknown state ($\boldsymbol{\beta}_t$) and its uncertainty over time in a state-space model. Hence we use Pólya-Gamma augmentation to make it tractable.

##### 5.4 Pólya–Gamma data augmentation

Following Polson *et al.* (2013) [8], we introduce a latent variable $\psi_i$ for each observation $y_i$, such that

$$\psi_i \sim \mathrm{PG}(n_i, z_i), \quad z_i = \mu + \phi(s_i)^\top \beta_{t_i}. \tag{12}$$

The Pólya–Gamma distribution PG$(b, c)$ is defined on $\psi > 0$ with density:

$$p(\psi \mid b, c) = \cosh^b\left(\frac{c}{2}\right) \frac{2^b}{\pi} \sum_{n=0}^{\infty} (-1)^n \frac{(n+b-1)!}{n!(b-1)!} \frac{2n+b}{\psi^{3/2}} \exp\left(-\frac{(2n+b)^2}{8\psi} - \frac{c^2\psi}{2}\right), \tag{13}$$

for parameters $b > 0$ and $c \in \mathbb{R}$ [8]. Although this infinite-series form is rarely evaluated directly, it provides a valid constructive definition and enables efficient sampling schemes. Importantly, the expectation of the PG distribution has a simple closed form:

$$\mathbb{E}[\psi \mid b, c] = \frac{b}{2|c|} \tanh\left(\frac{|c|}{2}\right), \tag{14}$$

with the limiting value $\mathbb{E}[\psi] = b/4$ as $|c| \to 0$. This property makes it well suited for Expectation–Maximization (EM) and variational algorithms, where only the first moment is required.

In our case, PG augmentation allows the binomial likelihood to be expressed as a Gaussian kernel conditional on $\psi_i$. Specifically, for the logistic likelihood term we have

$$p(y_i \mid z_i) \propto \exp\{\kappa_i z_i\} \int_0^\infty \exp\left(-\frac{\psi_i z_i^2}{2}\right) p(\psi_i)\, d\psi_i, \tag{15}$$

where $\kappa_i = y_i - n_i/2$. Conditioning on $\psi_i$ yields a Gaussian form in $z_i$:

$$p(y_i \mid \psi_i, z_i) \propto \exp\left(-\frac{\psi_i}{2}\left(z_i - \tfrac{\kappa_i}{\psi_i}\right)^2\right). \tag{16}$$

Thus, given the current latent values $\psi_i$, the observation model becomes conditionally Gaussian:

$$r_i = \frac{\kappa_i}{\psi_i} = \mu + \phi(s_i)^\top \beta_{t_i} + \varepsilon_i, \quad \varepsilon_i \sim \mathcal{N}(0, \psi_i^{-1}). \tag{17}$$

Here, $r_i = \kappa_i/\psi_i$ can be interpreted as a set of pseudo-observations. The Pólya–Gamma transformation replaces the original non-Gaussian likelihood with a conditionally Gaussian model over these pseudo-observations.

This formulation allows Kalman filtering and smoothing to be applied directly in the feature space, replacing the original non-Gaussian likelihood with a Gaussian pseudo-observation model.

##### 5.5 Approximate inference via EM with Kalman filtering and smoothing

We perform approximate inference using an EM algorithm.

**E-step**

In the E-step, we compute the expected values of the latent PG variables $\psi_i$ given the current estimates of the linear predictors $z_i = \mu + \phi(s_i)^\top \beta_{t_i}$:

$$\mathbb{E}[\psi_i \mid z_i] = \frac{n_i}{2|z_i|} \tanh\left(\frac{|z_i|}{2}\right), \tag{18}$$

with the limiting value $\mathbb{E}[\psi_i] = n_i/4$ as $|z_i| \to 0$. This result leverages the simple first moment of the PG distribution, despite its unwieldy density function.

These expectations define a set of observation-specific precisions $\Psi_t = \mathrm{diag}(\mathbb{E}[\psi_i])$ for all data points observed at time $t$.

**M-step**

Given these expected precisions, the conditional observation model becomes Gaussian:

$$r_t = \mu + \Phi_t \beta_t + \varepsilon_t, \quad \varepsilon_t \sim \mathcal{N}(0, \Psi_t^{-1}), \tag{19}$$

where $r_t = \Psi_t^{-1} \kappa_t$ and $\kappa_t = y_t - \frac{1}{2} n_t$. This pseudo-Gaussian model enables efficient updates of the latent feature coefficients $\beta_t$ using a Kalman filter (forward pass) followed by a Rauch–Tung–Striebel (RTS) smoother (backward pass).

**Kalman filtering and smoothing**

*Forward pass (Kalman filtering):*

The Kalman filter proceeds sequentially through time, propagating the mean and covariance of the latent coefficients given observations up to time $t$:

$$\text{Prediction:} \quad \beta_{t|t-1} = \beta_{t-1|t-1}, \tag{20}$$

$$P_{t|t-1} = P_{t-1|t-1} + Q, \quad Q = \tau^2 I_{2D}, \tag{21}$$

$$\text{Update:} \quad K_t = P_{t|t-1} \Phi_t^\top \left( \Phi_t P_{t|t-1} \Phi_t^\top + \Psi_t^{-1} \right)^{-1}, \tag{22}$$

$$\beta_{t|t} = \beta_{t|t-1} + K_t \big( r_t - \mu - \Phi_t \beta_{t|t-1} \big), \tag{23}$$

$$P_{t|t} = (I - K_t \Phi_t) P_{t|t-1}. \tag{24}$$

Here:

- $\beta_{t|t}$ and $P_{t|t}$ are the posterior mean and covariance of $\beta_t$ given data up to time $t$,
- $K_t$ is the Kalman gain matrix,
- $Q$ is the innovation covariance of the random walk.

*Backward pass (RTS smoothing):*

The filtering step provides estimates but only uses information up to the current time. To obtain posterior summaries that incorporate information from all time points, we perform a backward recursion:

$$J_t = P_{t|t} \, P_{t+1|t}^{-1}, \tag{25}$$

$$\beta_{t|T} = \beta_{t|t} + J_t (\beta_{t+1|T} - \beta_{t+1|t}), \tag{26}$$

$$P_{t|T} = P_{t|t} + J_t (P_{t+1|T} - P_{t+1|t}) J_t^\top, \tag{27}$$

where $\beta_{t|T}$ and $P_{t|T}$ denote the smoothed posterior mean and covariance of $\beta_t$ given all data up to time $T$.

**EM updates**

The E- and M-steps are alternated iteratively:

- E-step: compute $\mathbb{E}[\psi_i \mid z_i]$ from current $\beta_t$ estimates.
- M-step: run the Kalman filter and smoother to update $\beta_t, P_t$.

Each iteration refines the expected PG weights and the smoothed latent trajectories, yielding approximate maximum-likelihood estimates of the latent field.

Convergence of the EM algorithm can be assessed by monitoring the increase in the pseudo-Gaussian log-likelihood. After convergence of the EM algorithm, we obtain smoothed posterior means and covariances for the feature coefficients, $\beta_{t|T}, P_{t|T}$ for $t = 1 : T$. These summarize the estimated spatial-temporal dynamics of the latent logit-prevalence field in the reduced feature space.

##### 5.6 Reconstruction of the latent prevalence surface

Given the smoothed posterior mean and covariance of the latent coefficients, $\{\beta_{t|T}, P_{t|T}\}_{t=1}^T$, we obtain predictions for the latent prevalence surface by Monte Carlo sampling from the Gaussian posterior in feature space and transforming through the logistic link.

**1. Sample latent weights**

For each time $t$ and Monte Carlo draw $b = 1, \dots, B$:

$$\tilde{\beta}_t^{(b)} \sim \mathcal{N}(\beta_{t|T},\, P_{t|T}). \tag{28}$$

**2. Map to the logit scale and transform**

Calculate prevalence pointwise in $(s, t)$:

$$\tilde{z}^{(b)}(s,t) \;=\; \mu \;+\; \phi(s)^\top \tilde{\beta}^{(b)}(t), \qquad \tilde{p}^{(b)}(s,t) \;=\; \operatorname{logit}^{-1}\big(\tilde{z}^{(b)}(s,t)\big). \tag{29}$$

**3. Monte Carlo summaries**

$$\widehat{\mathbb{E}}\big[p(s,t)\big] = \frac{1}{B}\sum_{b=1}^{B} \tilde{p}^{(b)}(s,t), \tag{30}$$

$$\widehat{\mathrm{CI}}_{95\%}\big(p(s,t)\big) = \big[\,\tilde{p}^{(b)}_{2.5\%}(s,t),\; \tilde{p}^{(b)}_{97.5\%}(s,t)\,\big], \tag{31}$$

where $\tilde{p}^{(b)}_{q(s,t)}$ denotes the empirical $q$-th percentile across the Monte Carlo samples.

**4. Exceedance probability**

We calculate the exceedance probability, which in this case is the probability that the prevalence at a specific location $s$ and time $t$ was at or above a certain threshold:

$$\widehat{\Pr}\big(p(s,t) > p^\star \mid \mathbf{y}\big) = \frac{1}{B}\sum_{b=1}^{B} \mathbf{1}\Big\{\,\tilde{p}^{(b)}(s,t) > p^\star\,\Big\}, \tag{32}$$

where $\mathbf{1}\{\cdot\}$ is the indicator function and $p^\star$ is the chosen prevalence threshold. We set the prevalence threshold for the exceedance probability $p*$ at 5%.

##### 5.7 Variogram-based estimation of spatial and temporal correlation parameters

Spatial and temporal correlation parameters were estimated using empirical variogram analysis. Site-level prevalence estimates were transformed to the logit scale using a small-sample adjustment to stabilize variance. Geographic coordinates were projected into a locally defined Cartesian coordinate system using a Lambert azimuthal equal-area projection centred on the study region, allowing spatial separation to be expressed in kilometres.

Variogram analyses were conducted separately for two groups of molecular markers: (i) all *k13* mutations combined, and (ii) partner drug resistance markers combined (*crt* 76T and *mdr1* 86Y). Within each group, all available observations were pooled to compute a single spatiotemporal variogram, providing group-specific estimates of spatial and temporal correlation parameters.

For each marker group, a spatiotemporal variogram was computed using all available observation pairs. To isolate spatial dependence while minimizing confounding with temporal variation, the spatial variogram was extracted from pairs separated by approximately one year in time. The empirical spatial semivariogram was fitted using a Gaussian covariance model, with parameters estimated by weighted least squares, yielding an estimate of the spatial correlation length scale for each marker group.

Temporal variability was characterized using a temporal variogram constructed by restricting spatial separation to short distances and binning observation pairs by time lag. Assuming a first-order random-walk (RW1) temporal structure, the temporal semivariance was modeled as a linear function of time lag with the intercept constrained to zero. The slope of this relationship was estimated by weighted linear regression and used to derive the temporal variance parameter.

In subsequent spatiotemporal modelling, the estimated *k13*-specific spatial length scale was applied uniformly to all individual *k13* mutations, while the partner drug spatial length scale was applied to both *emph* 76T and *mdr1* 86Y.

We estimated the following length scale and temporal variability.

Table S4: Estimated spatial and temporal correlation parameters for antimalarial resistance mutations

| **Mutation** | $\ell_{\mathbf{km}}$ | $\tau^2$ |
|---|---|---|
| *k13* all validated and candidate combined | 57.3 | 0.28 |
| *crt* 76T and *mdr1* 86Y combined | 263.4 | 0.58 |

### 6 Downstream analysis of predicted prevalence

#### 6.1 Time-series analysis

Posterior predictions of mutation prevalence were obtained on a regular spatial grid as a four-dimensional array $\mathbf{P} \in \mathbb{R}^{n_x \times n_y \times T \times B}$, where $n_x$ and $n_y$ denote the spatial grid dimensions, $T$ the number of yearly time points, and $B$ the number of posterior draws. Each element $P_{i,j,t,b}$ represents a posterior draw of prevalence at spatial location $(i, j)$ and year $t$.

For each year $t$, the corresponding spatial slice $\mathbf{P}_{\cdot,\cdot,t,\cdot}$ was extracted and reshaped into a matrix with grid points as rows and posterior draws as columns. Grid points were assigned to administrative units using a precomputed spatial mapping. Let $\mathcal{S}_r$ denote the set of grid points belonging to Admin-1 region $r$.

For each region $r$, year $t$, and posterior draw $b$, regional mean prevalence was computed as

$$\bar{P}_{r,t,b} \;=\; \frac{1}{|\mathcal{S}_r|} \sum_{(i,j)\in\mathcal{S}_r} P_{i,j,t,b}. \tag{33}$$

This procedure yields a posterior distribution $\{\bar{P}_{r,t,b}\}_{b=1}^{B}$ for each region-year combination. Point estimates and uncertainty were summarized using posterior medians and corresponding 95% credible intervals across $b$, producing uncertainty-aware regional prevalence time series for downstream analysis and visualization.

#### 6.2 Area spread over time

For each molecular marker and year $t$, we define a fixed regular grid of locations $(x_k, y_k)$, indexed by grid cell $k$. From the spatiotemporal model, we obtain for each cell $k$ and year a posterior exceedance probability, as described above

$$\widehat{\Pr}\big(p(s,t) > p^\star \mid \mathbf{y}\big) = \frac{\sum_{b\in B} I(p^{(b)}(s,t) > p^\star)}{B}, \tag{34}$$

where $p^\star$ is the chosen prevalence threshold. We define the "spread" region via the exceedance probabilities. Specifically, for each year $t$, the set of grid cells that have an exceedance probability for the prevalence threshold, $p^\star$, is larger than the probability threshold $q_{\text{thr}} = 0.8$:

$$S_t = k : \widehat{\Pr}(t) \geq q_{\text{thr}}, \tag{35}$$

where $S_t$ is the set of grid cells where there is at least a $q_{\text{thr}}$ posterior probability that the prevalence exceeds the specified threshold $p^\star$. Each grid point is then buffered into a square polygon with side length equal to the grid spacing (here 5km), giving each cell an area $a_k$ in km$^2$. The total area of spread at year $t$, $A(t)$ is defined as the sum of the areas of all cells whose exceedance probability exceeds $q_{\text{thr}}$

$$A(t) = \sum_{k\in S_t} a_k. \tag{36}$$

#### References

[1] Covidence systematic review software, Veritas Health Innovation, Melbourne, Australia. Available at www.covidence.org.

[2] Junko Takata, Paul Sondo, Georgina S. Humphreys, Rebekah Burrow, Brittany Maguire, Mohammad S. Hossain, Debashish Das, Robert J. Commons, Ric N. Price, and Philippe J. Guerin. The WorldWide Antimalarial Resistance Network Clinical Trials Publication Library: A Live, Open-Access Database of Plasmodium Treatment Efficacy Trials. *The American Journal of Tropical Medicine and Hygiene*, 103(1):359–368, July 2020.

[3] MalariaGEN, Muzamil Mahdi Abdel Hamid, Mohamed Hassan Abdelraheem, Desmond Omane Acheampong, Ambroise Ahouidi, Mozam Ali, Jacob Almagro-Garcia, Alfred Amambua-Ngwa, Chanaki Amaratunga, Lucas Amenga-Etego, Ben Andagalu, Tim Anderson, Voahangy Andrianaranjaka, Ifeyinwa Aniebo, Enoch Aninagyei, Felix Ansah, Patrick O. Ansah, Tobias Apinjoh, Paulo Arnaldo, Elizabeth Ashley, Sarah Auburn, Gordon A. Awandare, Hampate Ba, Vito Baraka, Alyssa Barry, Philip Bejon, Gwladys I. Bertin, Maciej F. Boni, Steffen Borrmann, Teun Bousema, Marielle Bouyou-Akotet, Oralee Branch, Peter C. Bull, Huch Cheah, Keobouphaphone Chindavongsa, Thanat Chookajorn, Kesinee Chotivanich, Antoine Claessens, David J. Conway, Vladimir Corredor, Erin Courtier, Alister Craig, Umberto DAlessandro, Souleymane Dama, Nicholas Day, Brigitte Denis, Mehul Dhorda, Mahamadou Diakite, Abdoulaye Djimde, Christiane Dolecek, Arjen Dondorp, Seydou Doumbia, Chris Drakeley, Eleanor Drury, Patrick Duffy, Diego F. Echeverry, Thomas G. Egwang, Sonia Maria Mauricio Enosse, Berhanu Erko, Rick M. Fairhurst, Abdul Faiz, Caterina A. Fanello, Mark Fleharty, Matthew Forbes, Mark Fukuda, Dionicia Gamboa, Anita Ghansah, Lemu Golassa, Sonia Goncalves, G. L. Abby Harrison, Sara Anne Healy, Jason A. Hendry, Anastasia Hernandez-Koutoucheva, Tran Tinh Hien, Catherine A. Hill, Francis Hombhanje, Amanda Hott, Ye Htut, Mazza Hussein, Mallika Imwong, Deus Ishengoma, Scott A. Jackson, Chris G. Jacob, Julia Jeans, Kimberly J. Johnson, Claire Kamaliddin, Edwin Kamau, Jon Keatley, Theerarat Kochakarn, Drissa S. Konate, Abibatou Konate, Aminatou Kone, Dominic P. Kwiatkowski, Myat P. Kyaw, Dennis Kyle, Mara Lawniczak, Samuel K. Lee, Martha Lemnge, Pharath Lim, Chanthap Lon, Kovana M. Loua, Celine I. Mandara, Jutta Marfurt, Kevin Marsh, Richard James Maude, Mayfong Mayxay, Oumou Maiga-Ascofare, Olivo Miotto, Toshihiro Mita, Victor Mobegi, Abdelrahim Osman Mohamed, Olugbenga A. Mokuolu, Jaqui Montgomery, Collins Misita Moranga, Ivo Mueller, Kathryn Murie, Paul N. Newton, Thang Ngo Duc, Thuy Nguyen, Thuy-Nhien Nguyen, Tuyen Nguyen Thi Kim, Hong Nguyen Van, Harald Noedl, Francois Nosten, Rintis Noviyanti, Vincent Ntui-Njock Ntui, Alexis Nzila, Lynette Isabella Ochola-Oyier, Harold Ocholla, Abraham Oduro, Irene Omedo, Marie A. Onyamboko, Jean-Bosco Ouedraogo, Kolapo Oyebola, Wellington Aghoghovwia Oyibo, Richard Pearson, Norbert Peshu, Aung P. Phyo, Christopher V. Plowe, Ric N. Price, Sasithon Pukrittayakamee, Huynh Hong Quang, Milijaona Randrianarivelojosia, Julian C. Rayner, Pascal Ringwald, Anna Rosanas-Urgell, Eduard Rovira-Vallbona, Valentin Ruano-Rubio, Lastenia Ruiz, David Saunders, Alex Shayo, Peter Siba, Victoria J. Simpson, Mahamadou S. Sissoko, Christen Smith, Xin-Zhuan Su, Colin Sutherland, Shannon Takala-Harrison, Arthur Talman, Livingstone Tavul, Ngo Viet Thanh, Vandana Thathy, Aung Myint Thu, Mahamoudou Toure, Antoinette Tshefu, Federica Verra, Joseph Vinetz, Thomas E. Wellems, Jason Wendler, Nicholas J. White, Georgia Whitton, William Yavo, and Rob W. van der Pluijm. Pf7: an open dataset of Plasmodium falciparum genome variation in 20,000 worldwide samples. *Wellcome Open Research*, 8:22, 2023.

[4] World Health Organization. WHO malaria threats map: tracking biological challenges to malaria control and elimination., 2017.

[5] Ali Rahimi and Benjamin Recht. Random Features for Large-Scale Kernel Machines. In *Neural Information Processing Systems*, 2007.

[6] MATTHIAS SEEGER. GAUSSIAN PROCESSES FOR MACHINE LEARNING. *International Journal of Neural Systems*, 14(02):69–106, 2004. _eprint: https://doi.org/10.1142/S0129065704001899.

[7] S. Bochner. Monotone Funktionen, Stieltjessche Integrale und harmonische Analyse. *Mathematische Annalen*, 108:378–410, 1933.

[8] Nicholas G. Polson, James G. Scott, and Jesse Windle. Bayesian Inference for Logistic Models Using Pólya–Gamma Latent Variables. *Journal of the American Statistical Association*, 108(504):1339–1349, 2013. Publisher: ASA Website _eprint: https://doi.org/10.1080/01621459.2013.829001.
