## Supplemental figures for "Mapping the prevalence of molecular markers of *Plasmodium falciparum* artemisinin partial resistance in Africa: a spatial-temporal modelling study"

December 22, 2025

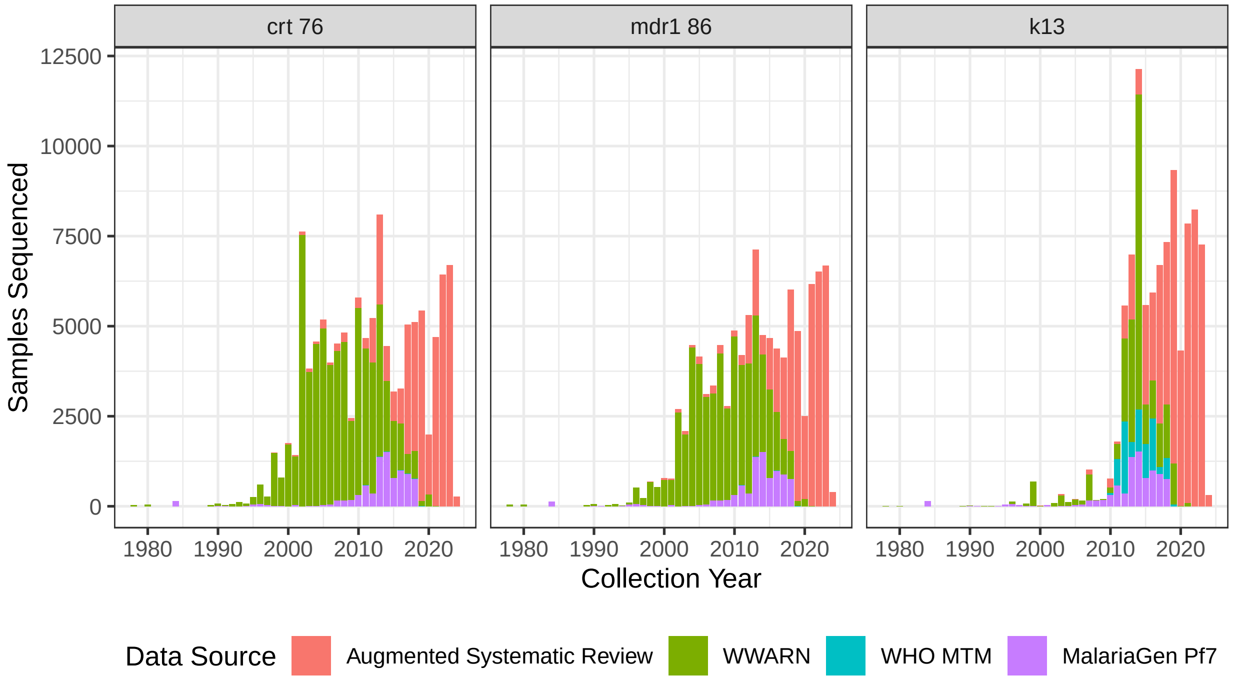


Figure S1: Number of samples in the combined dataset that were sequenced at each target position, from 1978 to 2024. For *k13* this includes all WHO validated and candidate ART-R mutation positions. Note that this is after ordered deduplication (Augmented Systematic Review ← WWARN ← WHO MTM ← MalariaGen Pf7).

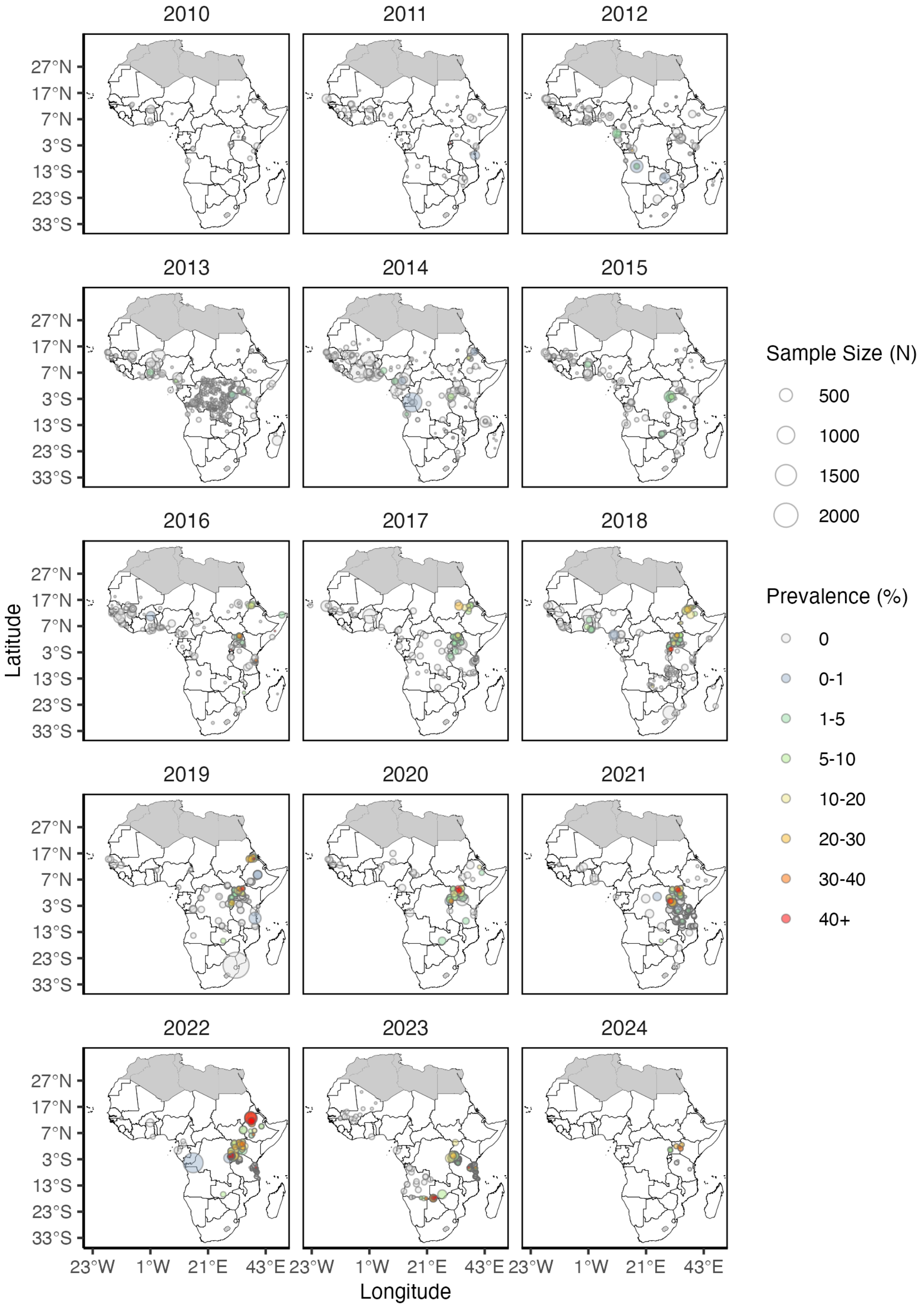


Figure S2: Observed prevalence of all WHO validated and candidate *k13* ART-R mutations combined across Africa from 2010 to 2024.

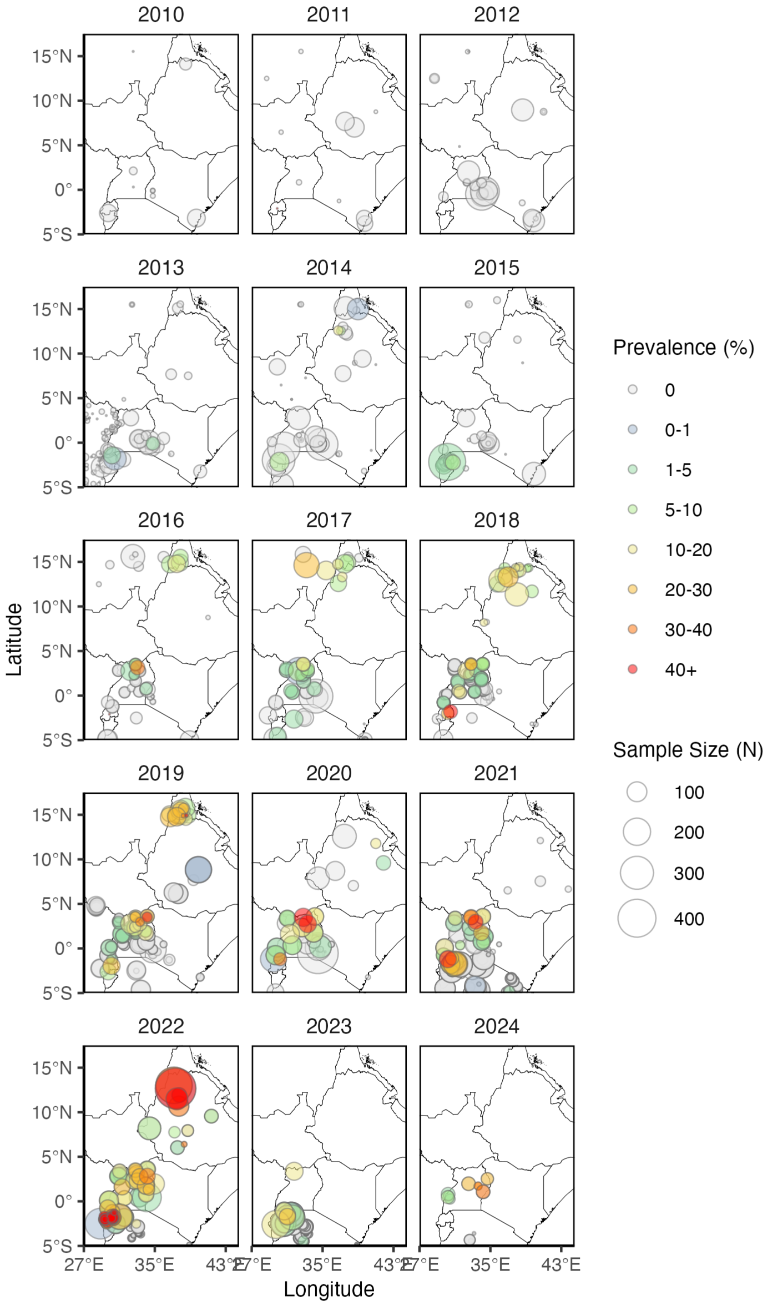


Figure S3: Observed prevalence of all WHO validated and candidate *k13* ART-R mutations combined across East Africa from 2010 to 2024.

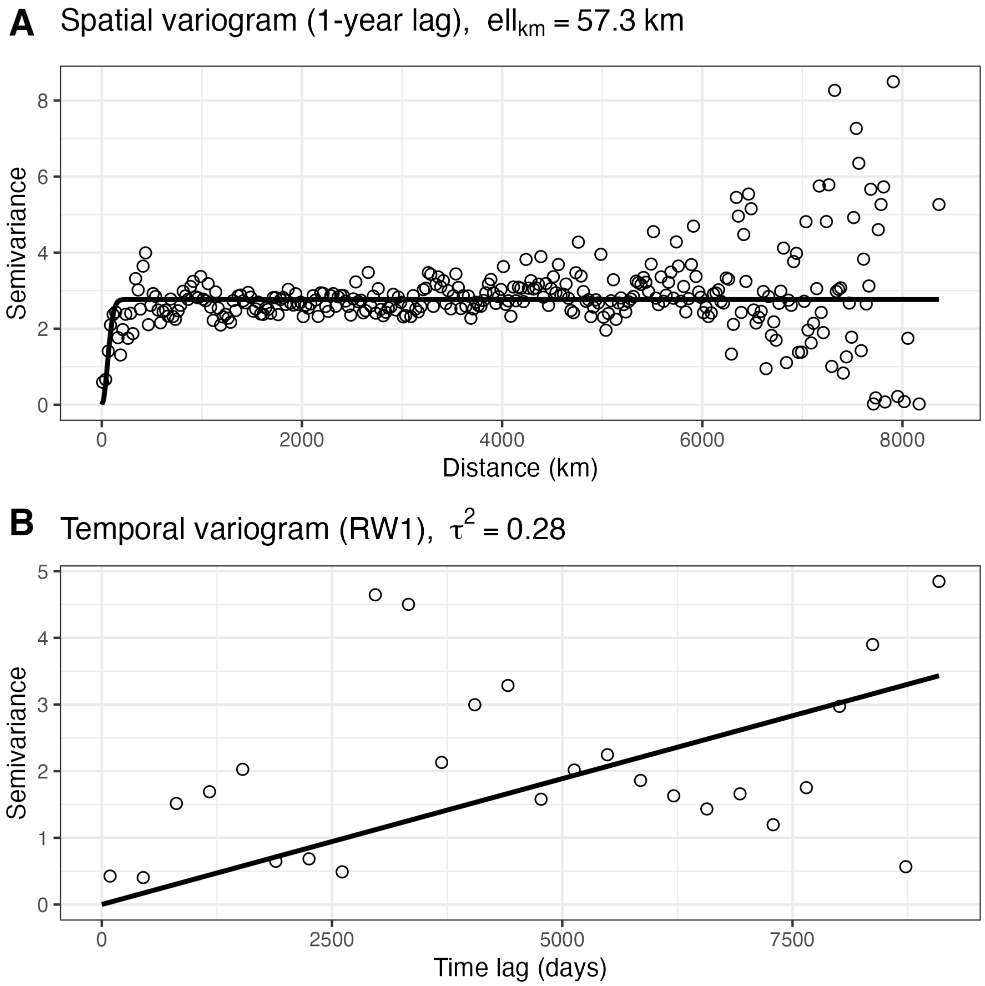


Figure S4: (A) Empirical spatial variogram for all validated and candidate *k13* mutations combined at an approximately one-year temporal lag, with a fitted Gaussian covariance model used to estimate the spatial correlation length scale. (B) Empirical temporal variogram with a fitted first-order random-walk model (intercept fixed at zero) used to estimate temporal variance. Points denote empirical semivariances and solid lines indicate fitted models.

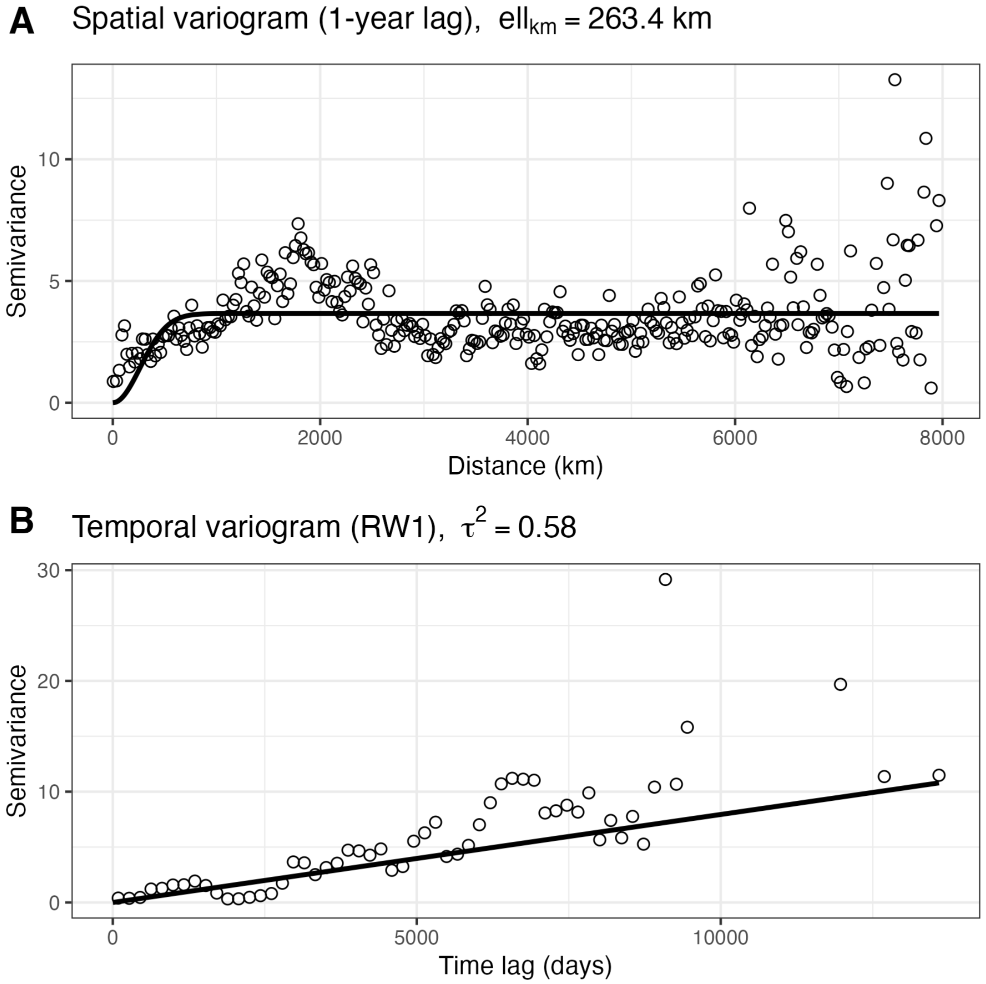


Figure S5: (A) Empirical spatial variogram for *crt* 76T and *mdr1* 86Y combined at an approximately one-year temporal lag, with a fitted Gaussian covariance model used to estimate the spatial correlation length scale. (B) Empirical temporal variogram with a fitted first-order random-walk model (intercept fixed at zero) used to estimate temporal variance. Points denote empirical semivariances and solid lines indicate fitted models.

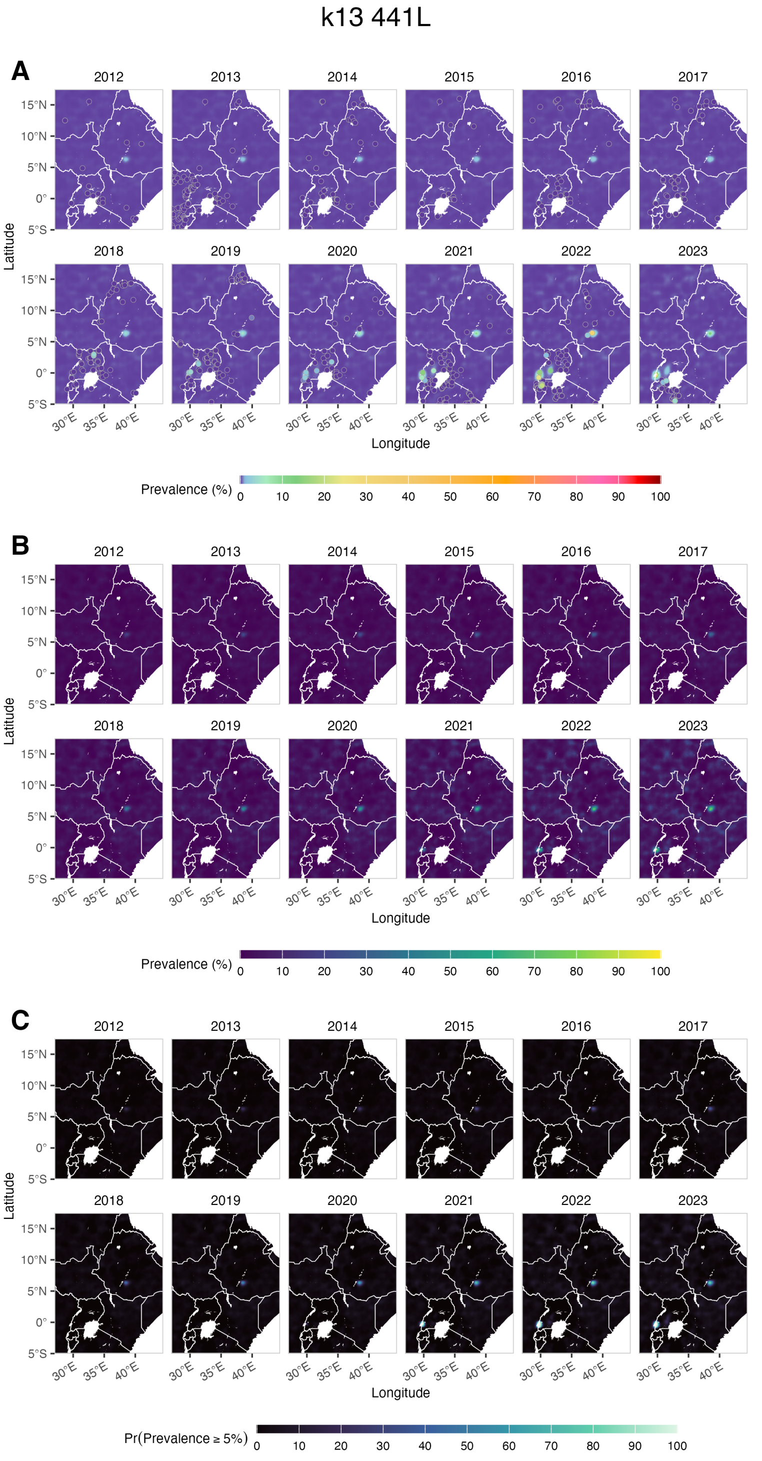


Figure S6: Predicted median prevalence surface with observed survey data overlaid (A), the width of the 95% credible interval (upper minus lower bound) representing predictive uncertainty (B), and the probability that prevalence exceeds 5% (C) for the 441L mutation in East Africa.

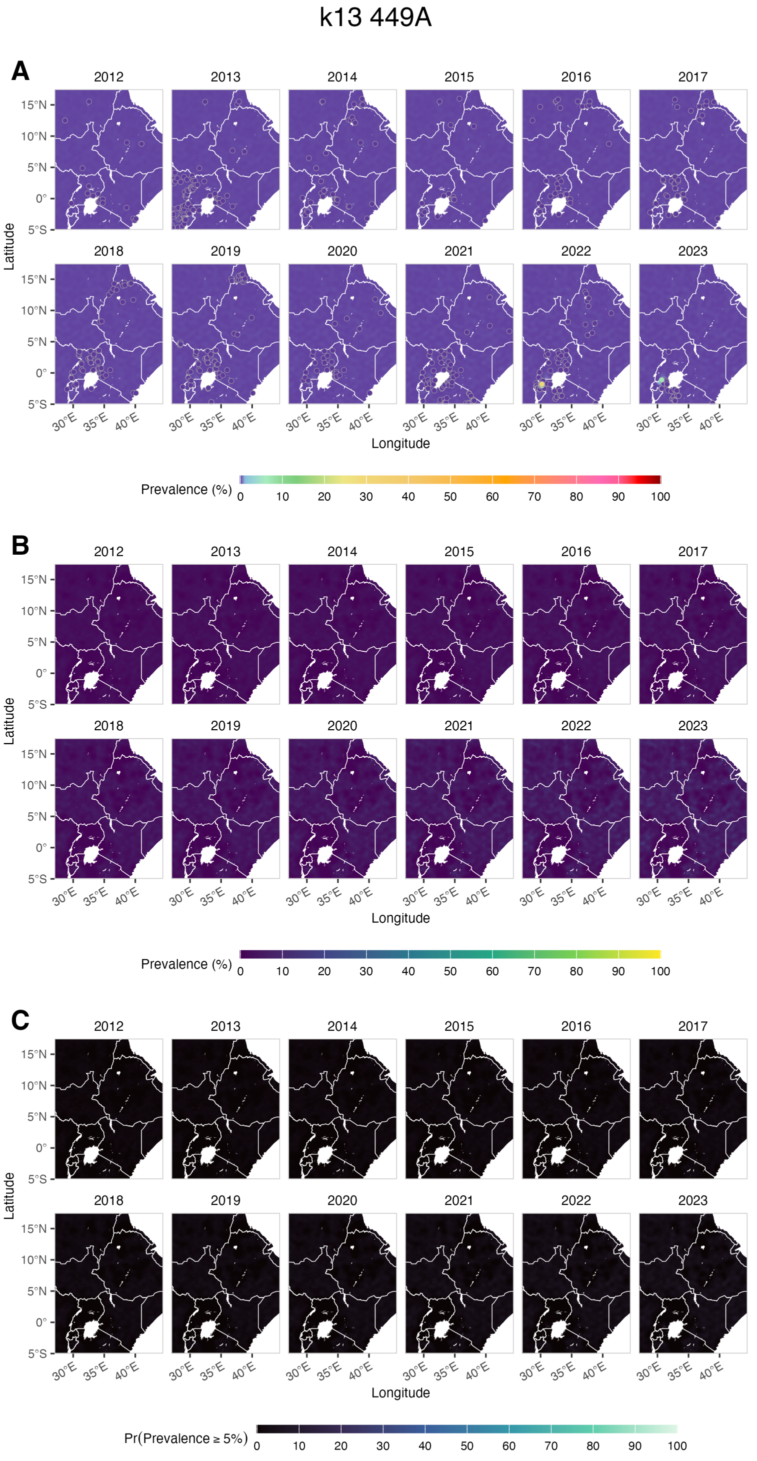


Figure S7: Predicted median prevalence surface with observed survey data overlaid (A), the width of the 95% credible interval (upper minus lower bound) representing predictive uncertainty (B), and the probability that prevalence exceeds 5% (C) for the 449A mutation in East Africa.

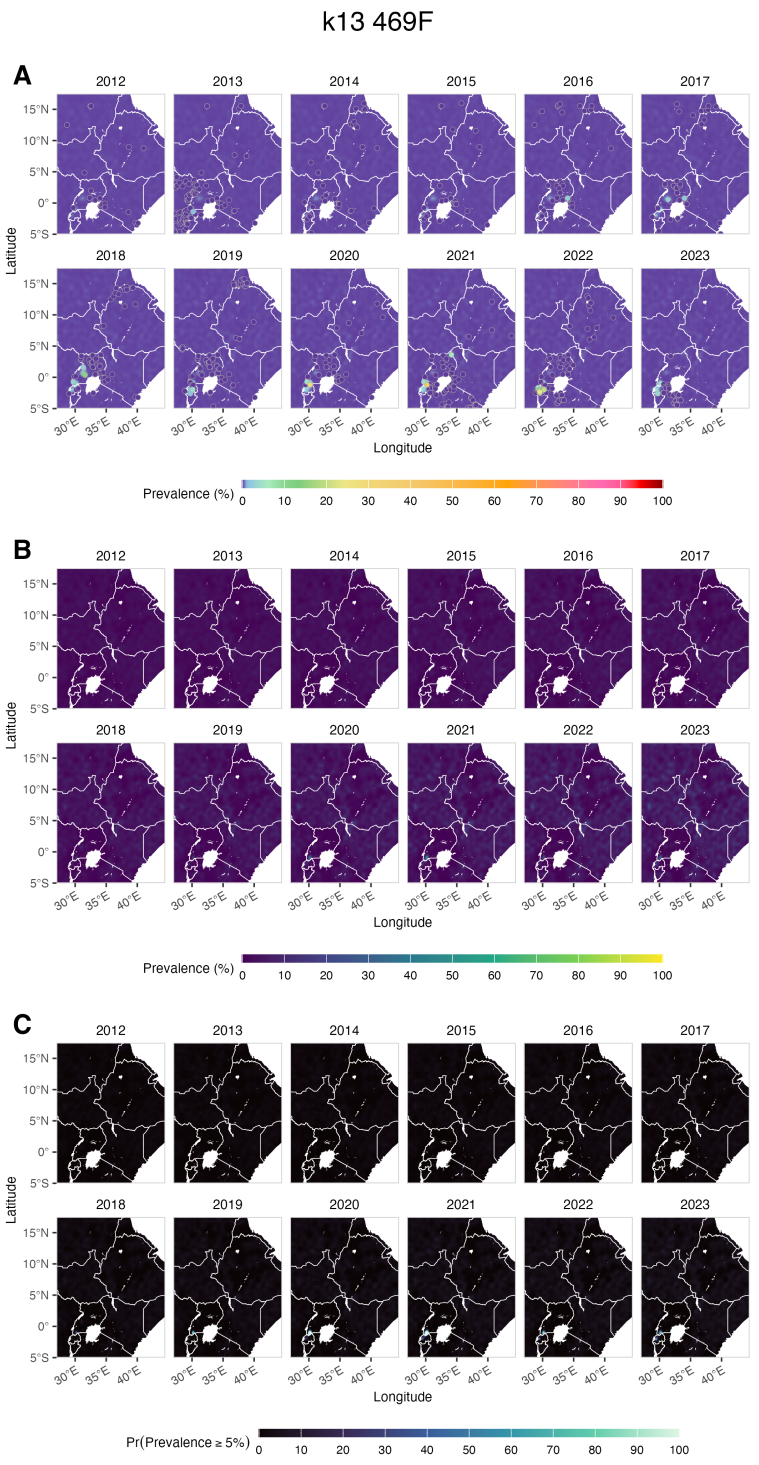


Figure S8: Predicted median prevalence surface with observed survey data overlaid (A), the width of the 95% credible interval (upper minus lower bound) representing predictive uncertainty (B), and the probability that prevalence exceeds 5% (C) for the 469F mutation in East Africa.

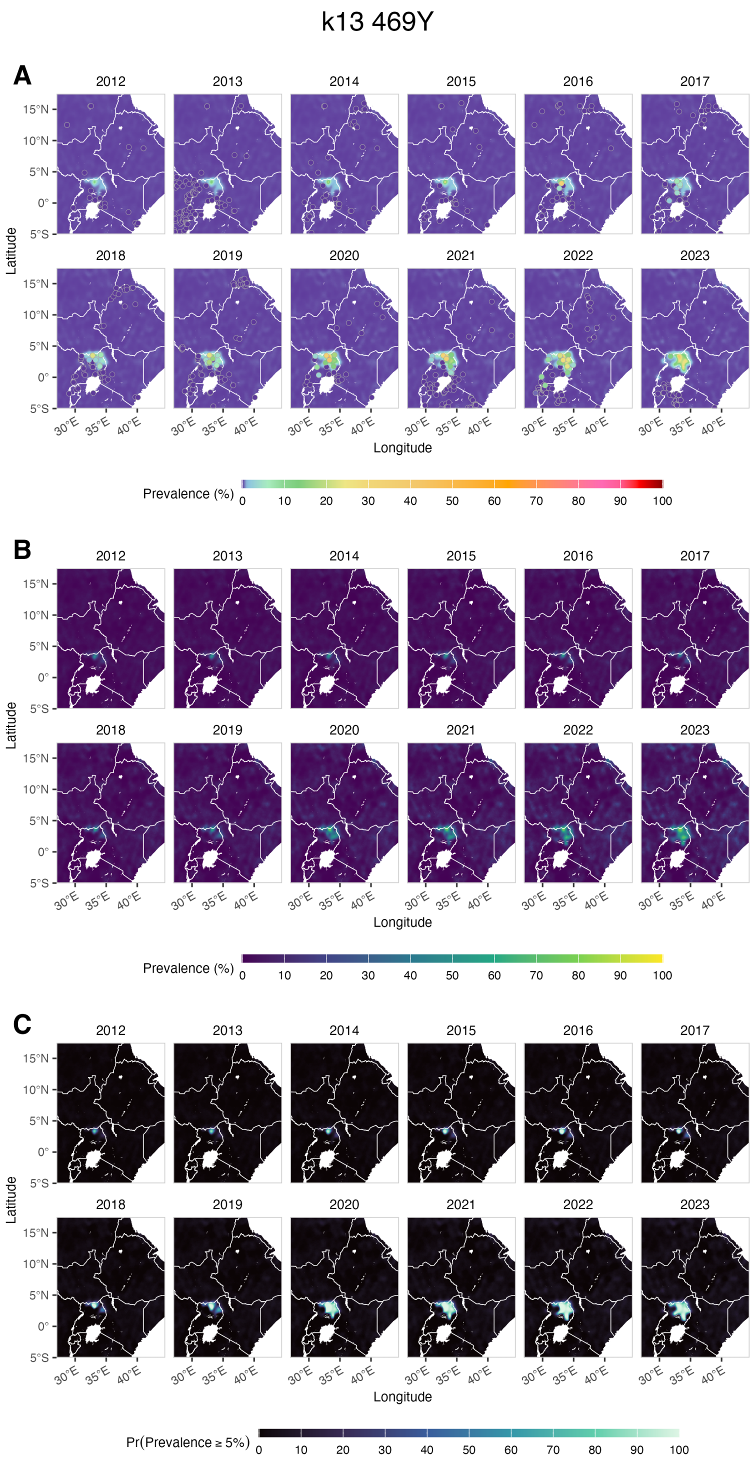


Figure S9: Predicted median prevalence surface with observed survey data overlaid (A), the width of the 95% credible interval (upper minus lower bound) representing predictive uncertainty (B), and the probability that prevalence exceeds 5% (C) for the 469Y mutation in East Africa.

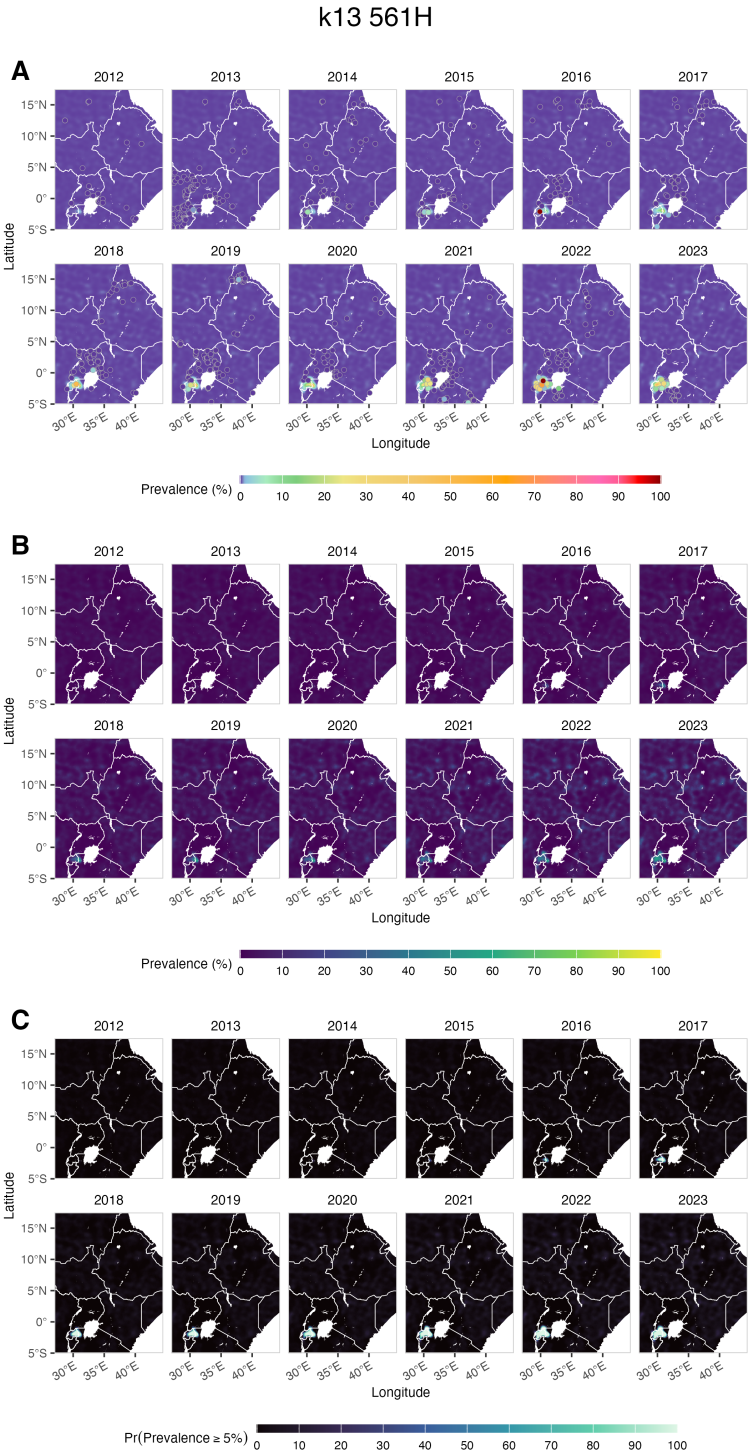


Figure S10: Predicted median prevalence surface with observed survey data overlaid (A), the width of the 95% credible interval (upper minus lower bound) representing predictive uncertainty (B), and the probability that prevalence exceeds 5% (C) for the 561H mutation in East Africa.

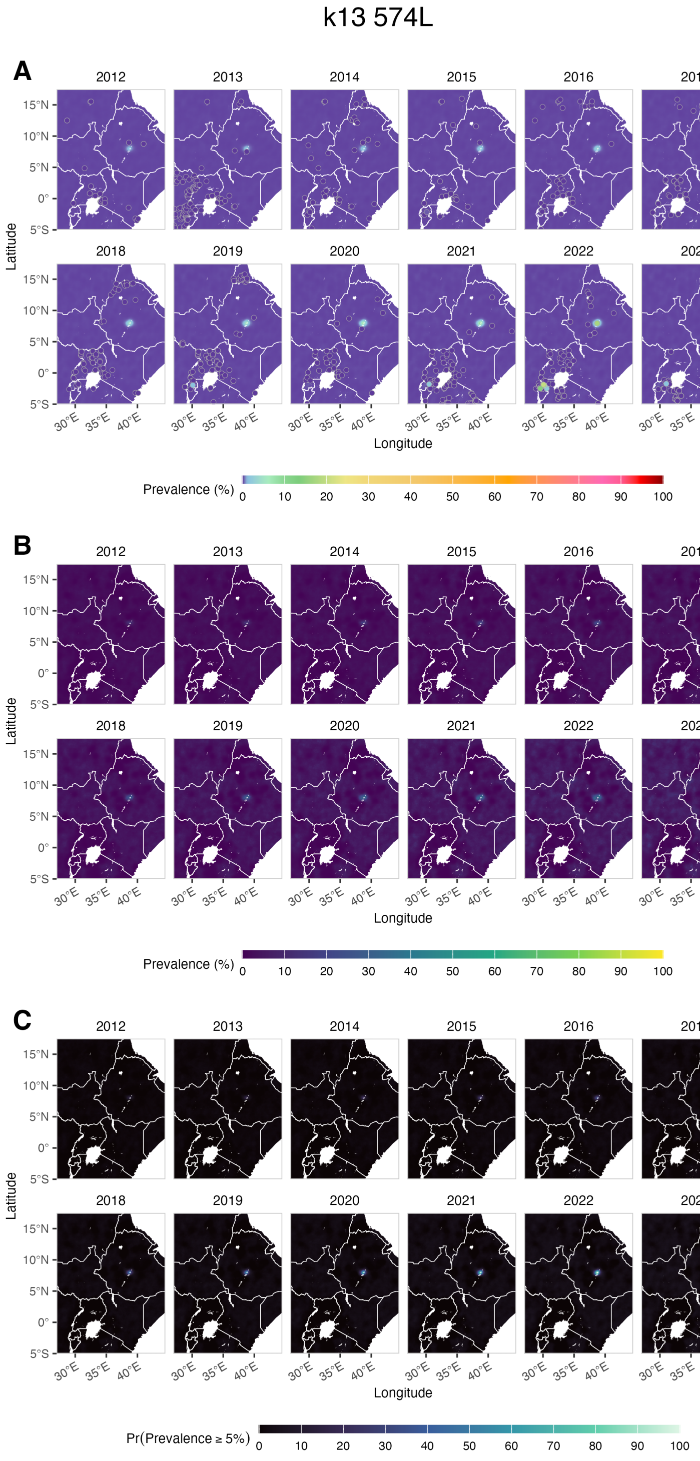


Figure S11: Predicted median prevalence surface with observed survey data overlaid (A), the width of the 95% credible interval (upper minus lower bound) representing predictive uncertainty (B), and the probability that prevalence exceeds 5% (C) for the 574L mutation in East Africa.

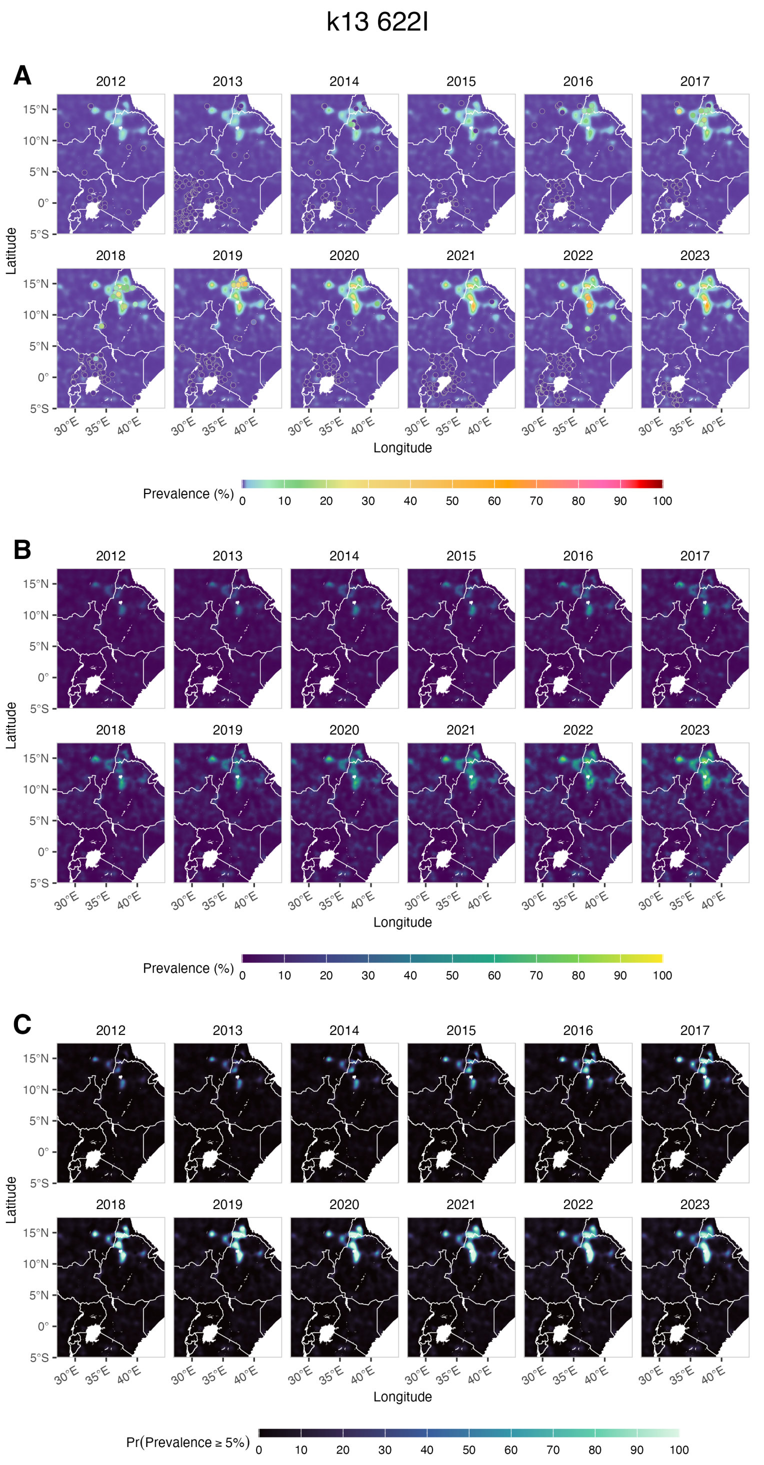


Figure S12: Predicted median prevalence surface with observed survey data overlaid (A), the width of the 95% credible interval (upper minus lower bound) representing predictive uncertainty (B), and the probability that prevalence exceeds 5% (C) for the 622I mutation in East Africa.

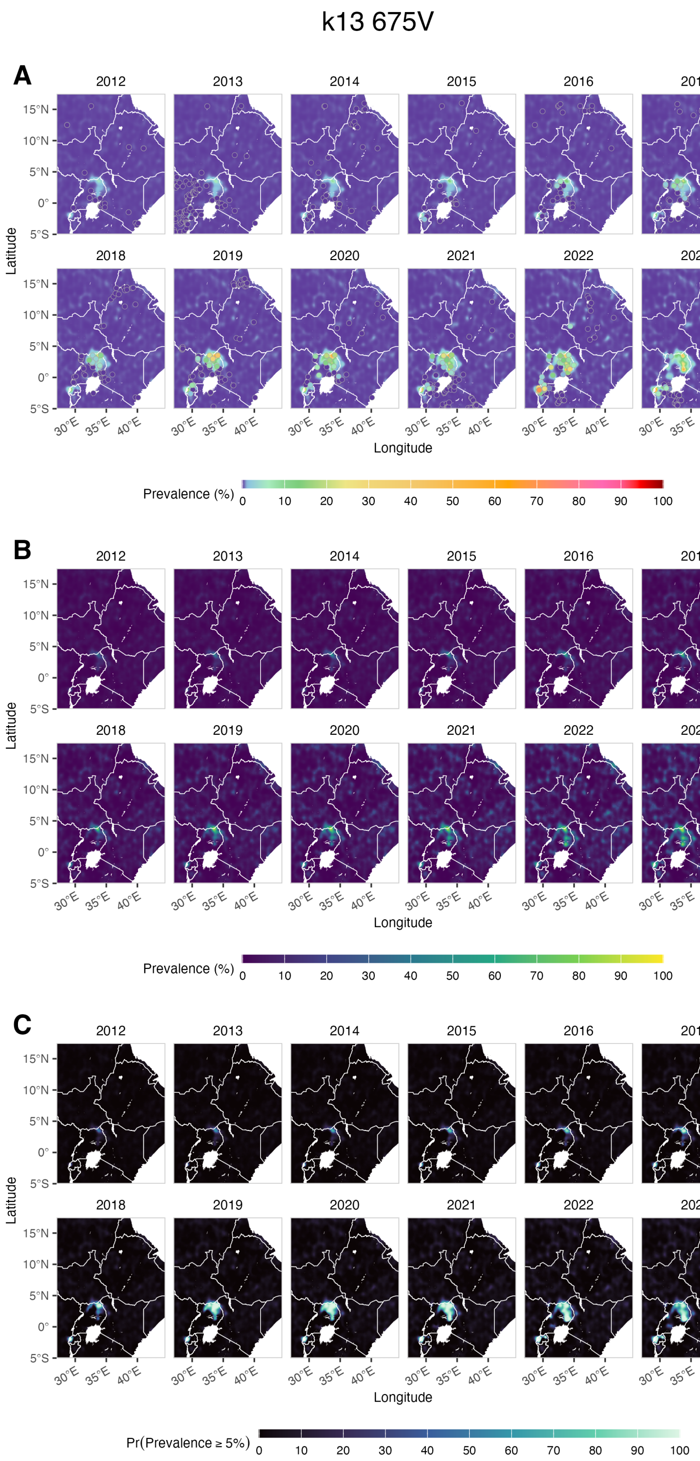


Figure S13: Predicted median prevalence surface with observed survey data overlaid (A), the width of the 95% credible interval (upper minus lower bound) representing predictive uncertainty (B), and the probability that prevalence exceeds 5% (C) for the 675V mutation in East Africa.

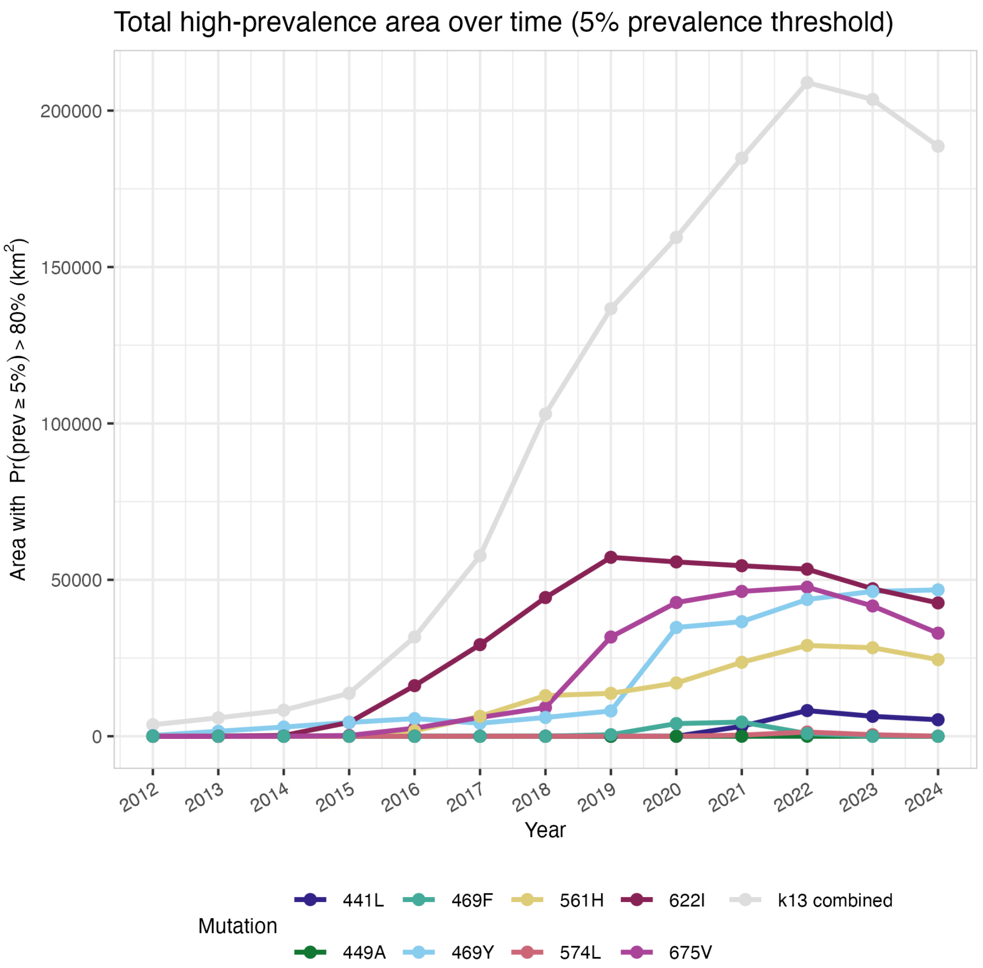


Figure S14: For each year, we count all pixels where the exceedance probability with threshold 5% (Pr(Prevalence > 5%)) is larger than 80%. The sum of those pixels gives a single total-area estimate per year for each mutation with data.

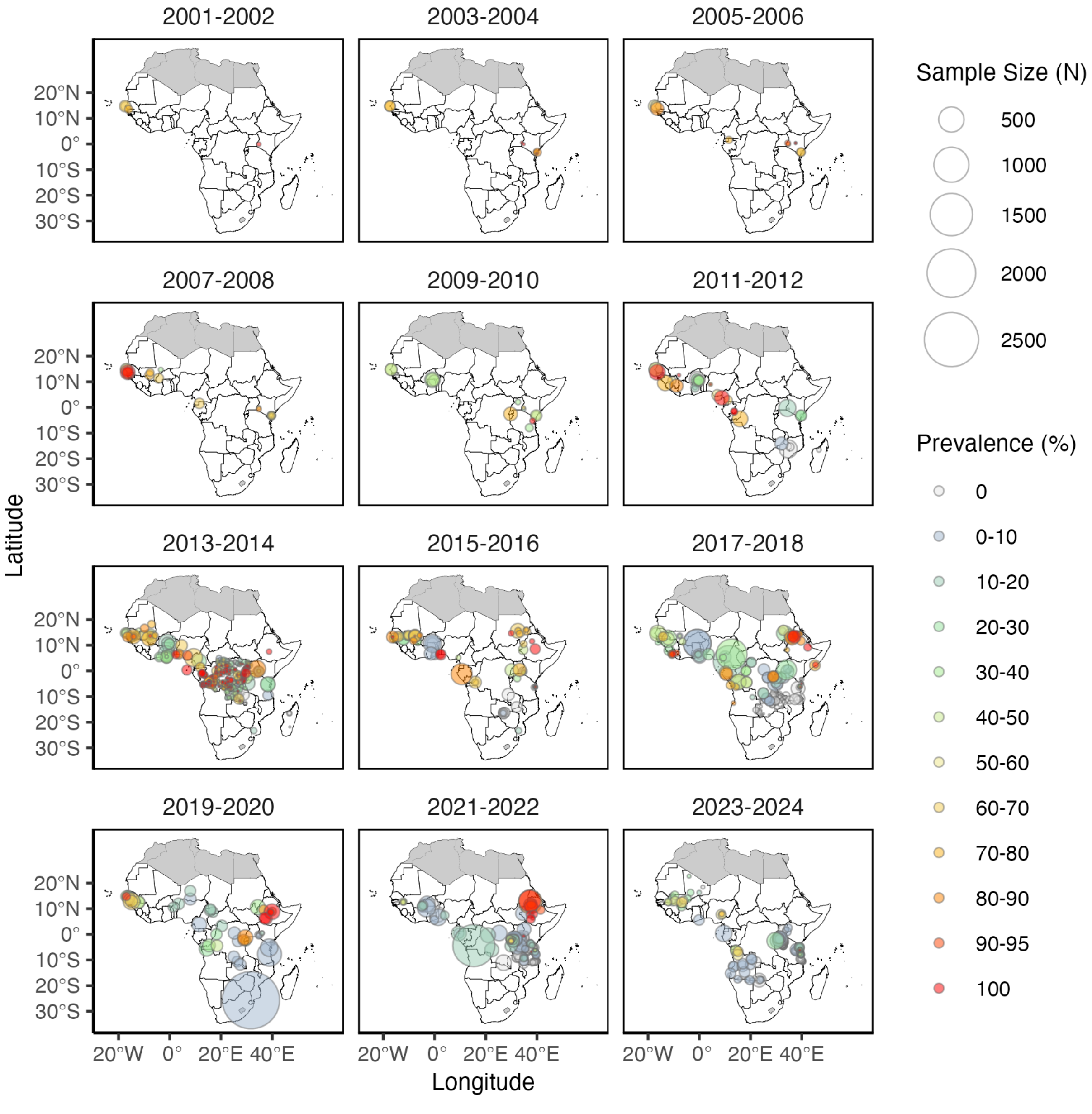


Figure S15: Map of all *crt* 76T datapoints from 2001 to 2024.

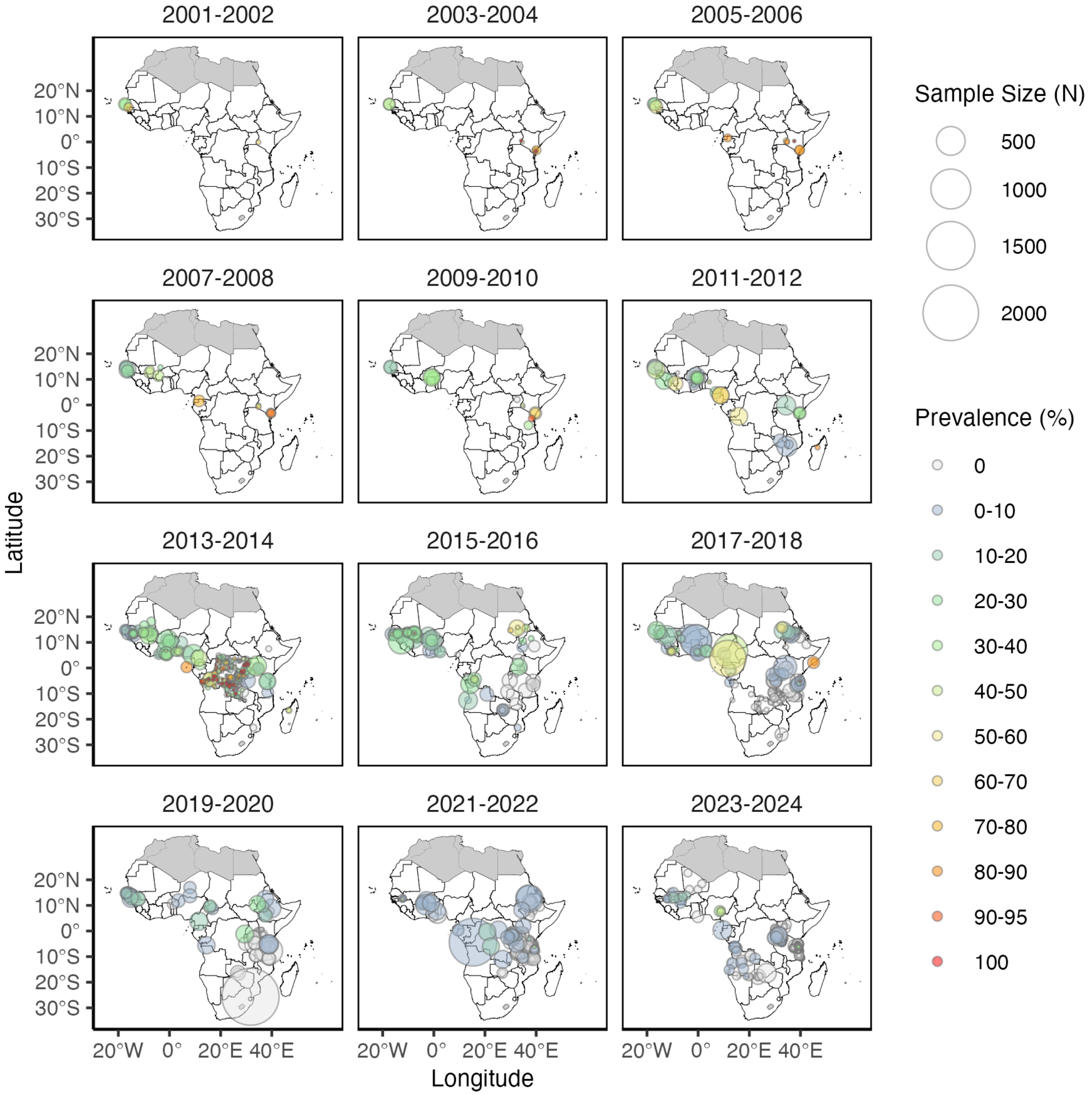


Figure S16: Map of all *mdr1* 86Y datapoints from 2001 to 2024.

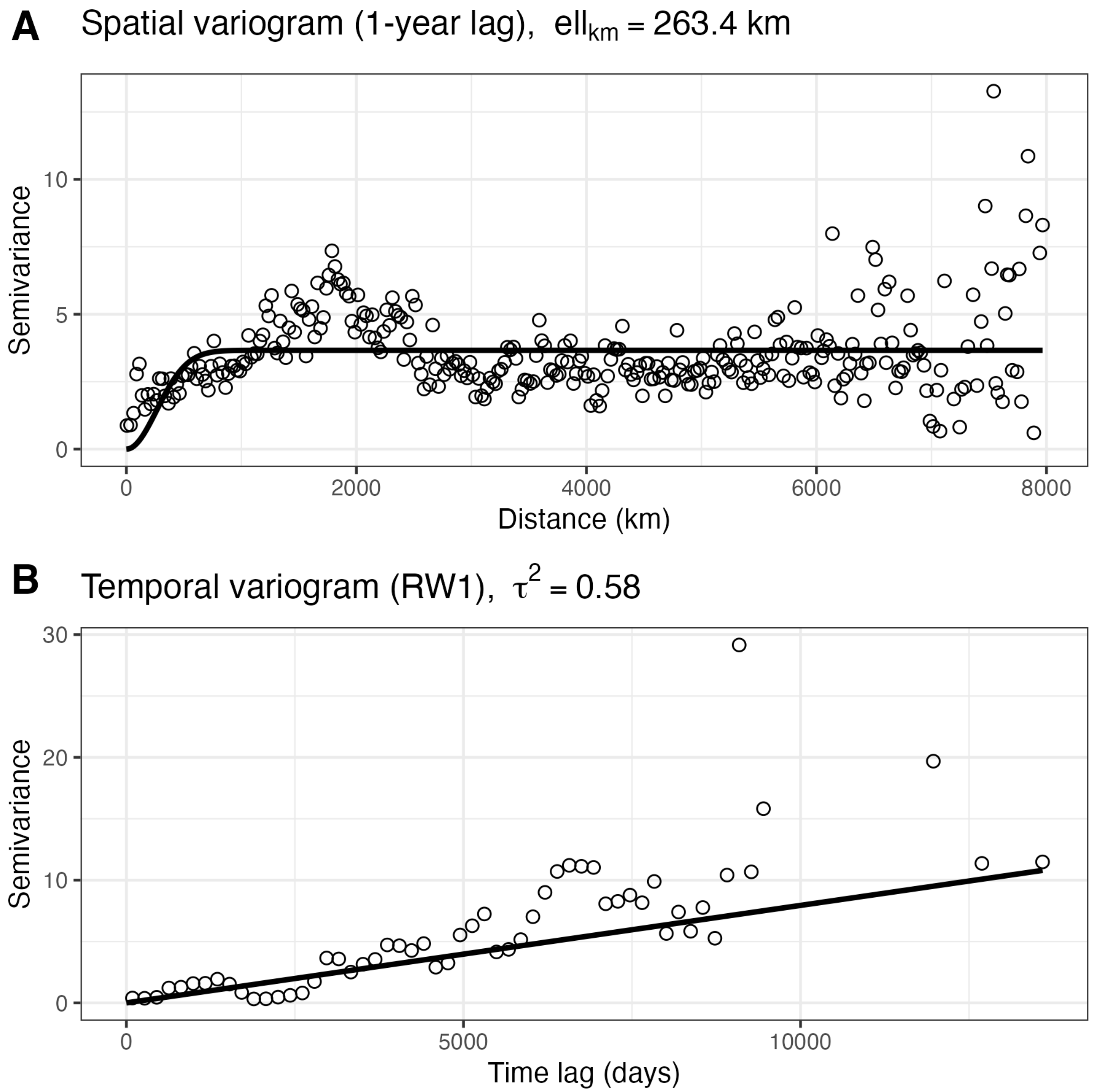


Figure S17: (A) Empirical spatial variogram for *crt* 76T and *mdr1* 86Y mutations combined at an approximately one-year temporal lag, with a fitted Gaussian covariance model used to estimate the spatial correlation length scale. (B) Empirical temporal variogram with a fitted first-order random-walk model (intercept fixed at zero) used to estimate temporal variance. Points denote empirical semivariances and solid lines indicate fitted models.

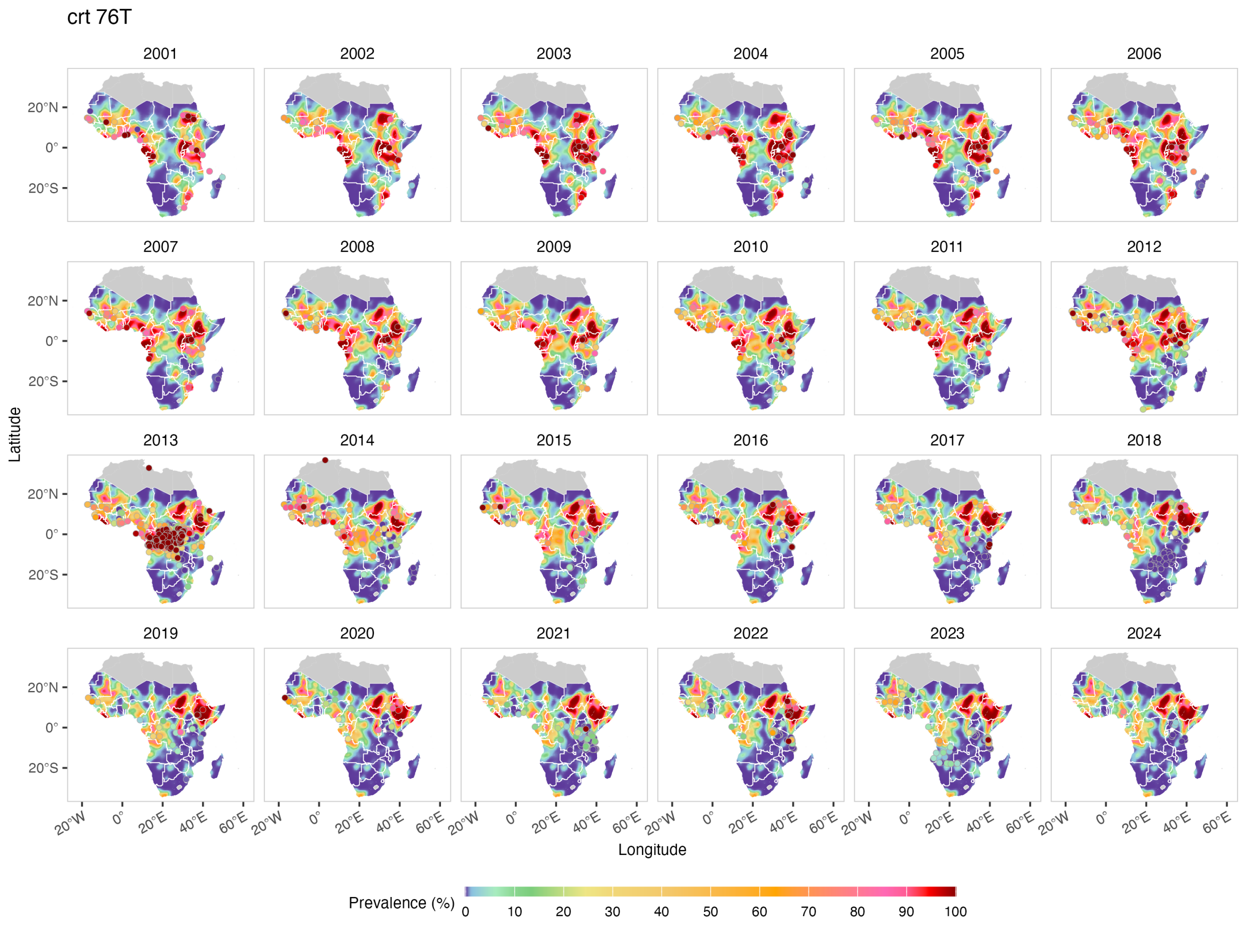


Figure S18: Predicted median prevalence of *crt* 76T across Africa from 2001 to 2024 with datapoints.

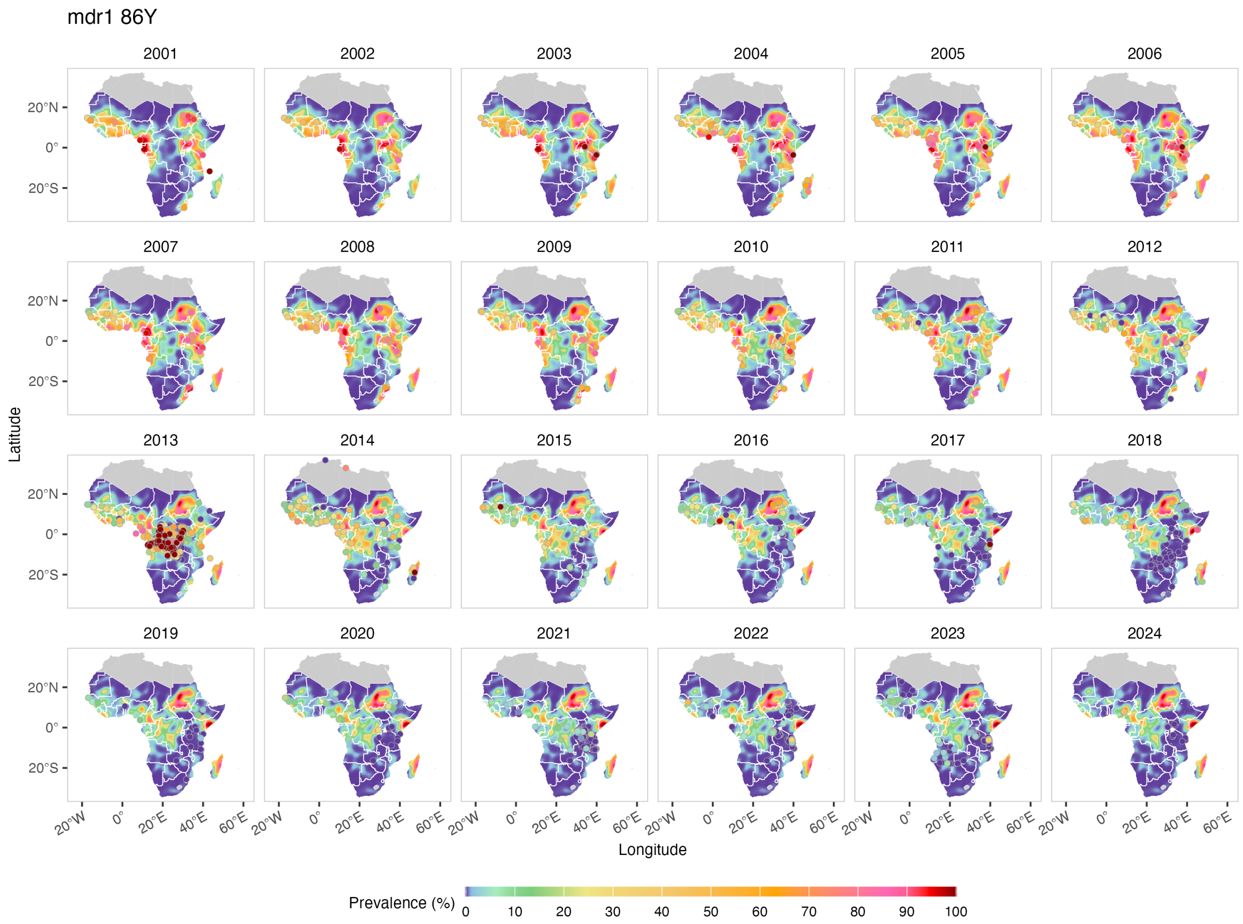


Figure S19: Predicted median prevalence of *mdr1* 86Y across Africa from 2001 to 2024 with datapoints.
